# Genetic dose-response modelling predicts drug mechanisms, dosing, and adverse events

**DOI:** 10.64898/2026.07.04.26357214

**Authors:** Luca Stefanucci, Daniel Considine, Ziying Ke, Joel Hellewell, Olivier Bakker, Marta P. Alcantara, Blagoje Soskic, Krista Freimann, Michael C. Turchin, Emily R. Holzinger, Joshua Chiou, Nikolina Nakic, Halit Ongen, Anna Lorenc, Maya Ghoussaini, Annalisa Buniello, Ian Dunham, Ellen M. McDonagh, Joseph C. Maranville, John A. Lees, Kaur Alasoo, David Ochoa, Yakov Tsepilov, Gosia Trynka

**Affiliations:** Wellcome Sanger institute, Wellcome Genome Campus, Hinxton, CB10 1SA, UK; Open Targets, Wellcome Genome Campus, Hinxton, CB10 1SA, UK; European Molecular Biology Laboratory, European Bioinformatics Institute, Wellcome Genome Campus, Hinxton, CB10 1SA, UK; Human Technopole, Viale Rita Levi-Montalcini 1, Milan, 20157, Italy; Institute of Computer Science, University of Tartu, Tartu, 51009, Estonia; Bristol Myers Squibb, Cambridge, MA, 2141, USA; Internal Medicine Research Unit, Pfizer Research and Development, 1 Portland St, Cambridge, MA, 02139, USA; Functional Genomics, Medicinal Science and Technology, GSK R&D, Stevenage, SG1 2NY, UK; Research Technologies, GSK, Heidelberg, Germany

## Abstract

Understanding how changes in gene function affect disease risk is central to drug development. Genetic variants are natural perturbations of gene activity and provide an opportunity to systematically model dose-response relationships between genes and phenotypes. Here, we present Variant-Informed Dose-Response Analysis (VIDRA), a computational framework that integrates trait-associated variants spanning a spectrum of allele frequencies and functional consequences within a hierarchical Bayesian regression model to systematically infer genetic dose-response relationships. Using 1,607,115 phenotypically associated germline variants available through Open Targets data (including common trait, rare disease, and gene burden associations), we systematically modelled how genetically driven alterations in gene function relate to disease risk, generating 148,350 dose-response-like gene-phenotype relationships across 7,681 phenotypes. Incorporating rare variant information alongside common variants resulted in a 7.23-fold increase in unique phenotypes, and a 51% increase in gene-disease pair associations. We calibrated our model against known drug targets to derive VIDRA Therapeutic Potential Score to rank genes based on their likelihood of succeeding as therapies, and identified 1,860 significant genes, 87% of which are currently therapeutically not targeted. VIDRA framework captures the direction and magnitude of gene-phenotype dependencies, enabling insights beyond target identification. Sixty-two percent of high-scoring targets are predicted to benefit from agonistic modulation, highlighting untapped potential for therapeutic activation. Furthermore, VIDRA framework extends to modelling relationships between genes, intermediate phenotypes, and diseases, enabling identification of biomarker-disease correlations. Applied to blood-cell and cardiovascular traits, VIDRA recovered known genetic links, suggesting a capacity to identify novel biomarker-disease relationships. Lastly, we observed a positive correlation by comparing VIDRA slopes with drug dose-response data from clinical trials, supporting the use of genetic data as a proxy for pharmacological titration. Together, VIDRA framework provides a generalizable, interpretable approach to inform multiple stages of drug development, from target prioritization and therapeutic direction of modulation prediction to biomarker identification, dose guidance, and safety risk assessment.

## Introduction

Human genetic studies are crucial in identifying causal genes that drive disease biology. In recent years, genetic associations have gained recognition as valuable tools for enhancing drug target prioritization, increasingly integrating into drug discovery workflows.^1–7^ Incorporating genetic evidence into drug development aims to address two major challenges: lack of efficacy and safety, which are among the leading causes of clinical trial failures.^8,9^ Lack of efficacy can stem from targeting genes that are merely correlated with, rather than causally involved in, disease biology. Retrospective analyses of clinical trials have shown that drug candidates with genetic support from common variant associations are at least twice as likely to gain regulatory approval, while drugs targeting Mendelian genes are seven times more likely to succeed through clinical trials into approved therapies.^5,10,11^ Additionally, GWAS signals driven by rare variants (MAF<0.1) with large effect sizes (OR>2) are four times more likely to succeed in drug program approvals than those based on common variants with small effects.^12^

With growing pharmacological and clinical interest, the number of gene-to-phenotype associations has surged since the completion of the Human Genome Project. This expansion has revealed genes with multiple independent alleles, each exerting distinct effects on protein activity or abundance. These variations, in turn, contribute to a spectrum of phenotypic alterations, ranging from imperceptible changes to severe outcomes. This gradation of phenotypic effects linked to multiple alleles is known as an allelic series.^13,14^ However, understanding the biological relevance of genetic associations requires interpreting diverse types of evidence, each potentially capturing different aspects of gene function.^15^ Common variant associations from GWAS predominantly identify common variants with modest effect sizes, often located in non-coding regulatory regions, where they may alter gene expression levels or splicing patterns. Some of these effects can be captured by quantitative trait loci (QTL) approaches, mapping the effects of variants on transcript (eQTLs)^16^ or protein (pQTLs)^17^ levels, that can be carried out across hundreds or thousands of individuals and across diversity of tissues^18^, cell types and cell states^19^, providing comprehensive maps of gene and protein expression regulation. Ultimately, such maps can provide linkage between GWAS associated signals and gene and protein levels through colocalization of associated signals.^20,21^ In contrast, rare coding variants, both single variant and those identified through gene burden tests, can directly implicate genes by demonstrating larger-effect sizes and typically are identified in the context of loss of gene function. These signals are often more interpretable but occur less frequently in the population and require large sample sizes or family pedigrees to be identified.

Depending on the gene, changes in function, whether through altered expression levels or activity, may have minimal impact or, conversely, lead to severe phenotypes. Some genes, however, are tolerant to partial or even complete loss of function, and in certain cases, such loss may confer disease protection with little to no adverse effects. For instance, *PCSK9* loss-of-function variants are associated with reductions of blood cholesterol levels,^22–24^ while gain-of-function variants can lead to familial forms of hypercholesterolemia.^25,26^ Ultimately, the knowledge of how the different magnitudes and directions of effects of genetic variants in PCSK9 protein, and the consequent effect on the human phenotype, led to the development of chemical PCSK9 inhibitors, such as Alirocumab and Evolocumab, to prevent cardiovascular events.^27,28^

A systematic and unbiased approach to studying gene function could leverage all naturally occurring variants that exert cis-effects on protein level or activity and human phenotypes, allowing for a quantitative assessment of their relationships. The use of broad sources of genetic variation information provide an unique coverage across a spectrum of protein activity and disease severity.^29^ Furthermore, many rare disease genes also harbour common variant associations, suggesting a continuum in the genetic architecture of diseases and their phenotypic co-occurrence.^30^ Integrating rare variants into allelic series modelling helps account for the effects of extreme gene function modulation that otherwise are missed if relying solely on common variants, as their effects are much more subtle. However, integrating these different sources of evidence into a single statistical framework remains challenging, particularly due to differences in effect size scales on traits and gene function. Developing models that can harmonise evidence across both coding and non-coding variants is essential for improving our ability to infer causal mechanisms and prioritise the most effective therapeutic targets.

Here, we present the Variant-Informed Dose-Response Analysis framework (VIDRA), which applies Mendelian randomization (MR) like approach to position genetics as a central tool for developing new therapeutic hypotheses. Following MR instrumental variable logic, genetic variants serve as instruments for gene function: variants are associated with both a molecular exposure (gene expression or protein activity) and a disease outcome, and the ratio of their effects provides an estimate of the causal dependency of phenotype on gene function. The VIDRA framework integrates multiple classes of genetic evidence within a hierarchical Bayesian regression model to estimate dose-response-like relationships between gene function and phenotypic traits. This continuum of genetic perturbations, from common variants with modest effects on gene expression to rare coding variants that severely disrupt protein function, is conceptually analogous to a pharmacological dose-response curve, in which graded modulation of a target produces proportionate phenotypic effects. We exploit this analogy explicitly: throughout this work, ‘dose’ refers to the genetically inferred level of protein activity perturbation, not a literal drug concentration. By the same principle, the ‘response’ refers to the phenotypic changes observed at a given level of genetically inferred perturbation. We systematically modeled the dose-response relationships between 18,838 genes and 7,681 phenotypes using 1,607,115 independent genetic variants. We used the Open Targets Platform as a source of variant information, which incorporates data from GWAS, QTL studies, and ClinVar, alongside loss-of-function variant gene burden results from the AZ PheWas (https://azphewas.com/).^7,31–34^ We used a MR-derived slope as a quantitative estimate of phenotypic changes based on genetic variation.^1,2,35^ We evaluated the performance of our approach and showed that VIDRA outperforms established tools in a retrospective analysis of successful drugs, demonstrating greater sensitivity in identifying potential drug targets. Beyond target prioritisation, VIDRA provided estimates of a drug’s therapeutic direction of modulation, whether it should function as an inhibitor or an activator, by analyzing if the disease-associated alleles lead to a gain or loss of function. Furthermore, we demonstrate how VIDRA models genetically inferred dose-response relationships akin to those used in drug development, which can aid in determining safe and effective drug doses that define the boundaries of actionable and tolerable gene modulation.^14^ Our findings broaden the role of genetics in early-stage drug discovery, extending beyond target prioritization to systematically informing therapeutic direction of modulation, efficacy, and potential adverse effects.

## Results

### Construction of genetic dose-response relationships using the VIDRA framework

#### Overview of the method

VIDRA is a computational framework that integrates multiple classes of genetic evidence and applies a hierarchical Bayesian regression model to estimate dose-response relationships between gene function and phenotypic traits. Throughout this manuscript, we refer to VIDRA as the framework when discussing its overall scope and to the VIDRA Bayesian model when referring specifically to its regression component. The VIDRA framework systematically infers genetic dose-response relationships between gene function alteration and disease risk by integrating heterogeneous classes of genetic evidence within a hierarchical Bayesian regression model. To approximate the effects of genetic variants on protein function we took the following approach (**Fig. 1**): (i) for unique non-coding variants (n = 35,760), our model uses disease GWAS colocalisation^20^ with molecular QTLs (eQTL n = 34,753; and pQTL n = 2,103), to approximate the effects of a non-coding variants on a transcript or protein expression; and (ii) for coding variants without QTL effects (GWAS-coding=26,665, AZ-rare=3,418, and ClinVar=1,549,806), we rely on an aggregated functional score from *in-silico* tools (such as VEP or CADD, see **Methods**). Utilisation of rare variant information resulted in a 7.23-fold increase in distinct phenotypes, and a 51% increase in unique gene-disease pairs, when compared to use of common GWAS variant information alone. When available, gene burden test results were used as priors for the intercept, approximating the baseline phenotype value when protein function is effectively absent (i.e., close to null). We observe that the aggregated variant effect prediction scores captured the effects on gene function when compared to experimental data from saturation genome editing studies (Pearson’s R=0.48, p-value=1×10^-227^; **Supplementary Fig. 1**). The phenotypes used in VIDRA framework encompass the full spectrum of human traits catalogued in Open Targets, including common complex diseases, rare Mendelian disorders, quantitative physiological measurements, and molecular traits such as circulating protein and metabolite levels, all unified under Experimental Factor Ontology (EFO) terms.

**Figure 1.**
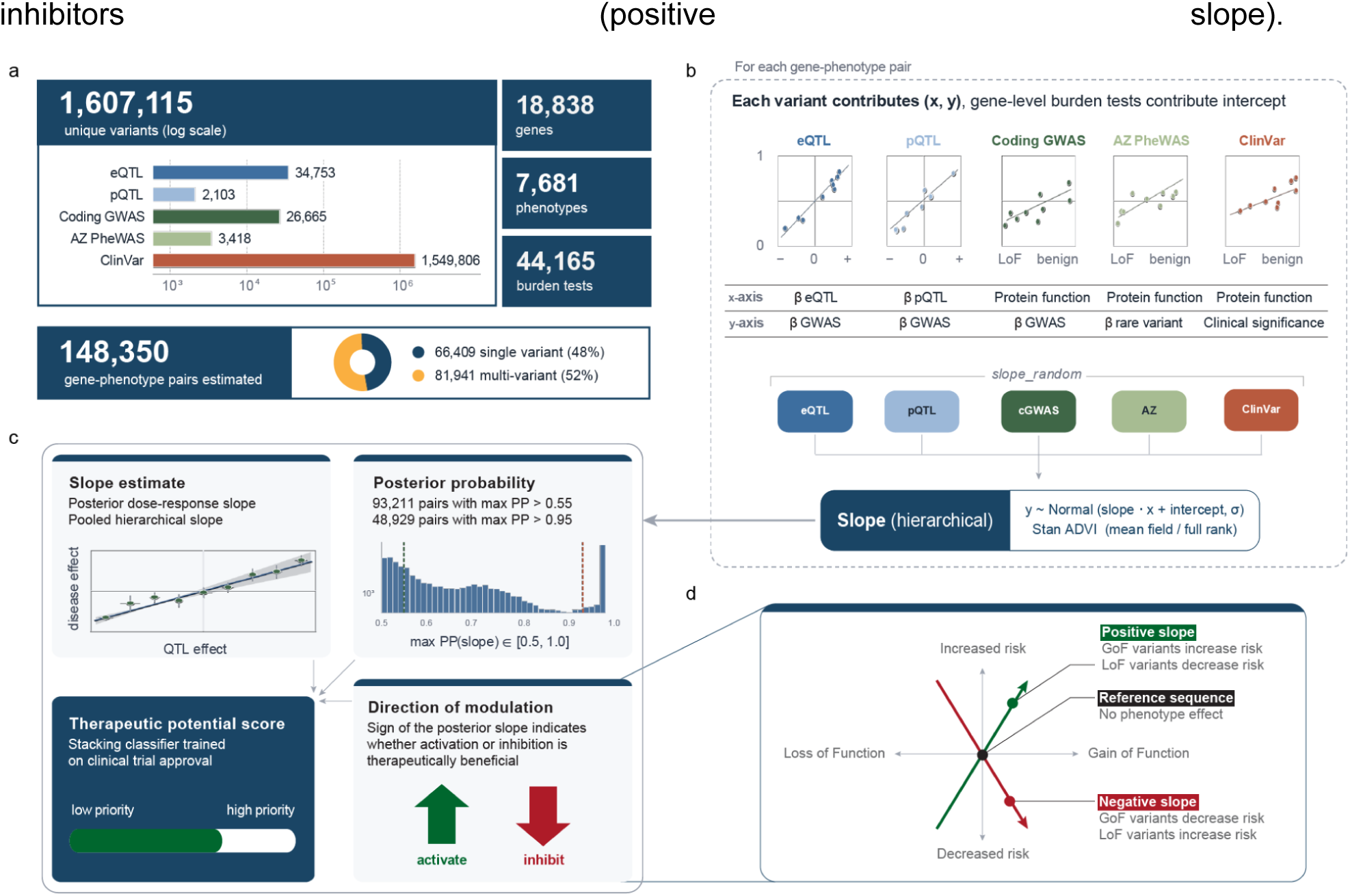
Overview of the VIDRA framework. **(a)** Data inputs. VIDRA integrates five complementary sources of genetic evidence: expression quantitative trait loci (eQTLs), protein quantitative trait loci, coding GWAS variants, AstraZeneca PheWAS variants, and ClinVar variants, together with gene-level rare variant burden tests. **(b)** Variant effect estimation. Each variant contributes a point to a dose-response model, where *y* represents disease effect and *x* represents either QTL effect size (eQTLs and pQTLs) or a protein function score (coding GWAS, AZ PheWAS, and ClinVar). Gene-level burden tests provide anchoring information for loss-of-function effects. Evidence is integrated using a hierarchical Bayesian model to estimate a gene–phenotype slope. **(c)** Outputs. For each gene–phenotype pair, VIDRA produces a posterior slope estimate, a posterior probability (PP) quantifying confidence in a non-zero slope, a Therapeutic Potential Score derived from a machine-learning classifier trained on approved drug targets, and a predicted direction of therapeutic modulation inferred from the slope sign. **(d)** Interpretation of slope direction. The VIDRA slope captures the relationship between protein activity and disease risk across the spectrum from loss-of-function (LoF) to gain-of-function (GoF). Positive slopes indicate that increased protein activity is associated with increased disease risk, whereas negative slopes indicate that increased protein activity is protective. Directionality is inferred from the combined genetic evidence and does not require explicit GoF or LoF annotation.

Briefly, the VIDRA Bayesian model fits a linear regression per gene-phenotype pair, yielding the VIDRA slope which captures the direction and magnitude of the dose-response relationship, representing the extent to which the phenotype of interest depends on the changes in relative protein function (**Methods**). To account for differences in data quality and biological interpretation, it uses a hierarchical model to estimate the slope distribution, accounting for different sources of genetic evidence. By modelling slopes as random effects within a hierarchical Bayesian regression model and using Student’s t-distribution to account for outliers, VIDRA effectively shares information across variant groups while controlling for noise. Slope estimates are inferred via Markov chain Monte Carlo sampling, and only gene-phenotype pairs with high posterior support (PP≥0.7 for ClinVar-only, PP≥0.55 for multi-source evidence) are retained for downstream analyses. The VIDRA slope quantifies how strongly a phenotypic trait depends on genetically-inferred protein activity; a positive slope indicates that increasing protein activity increases trait risk (an inhibitor target), while a negative slope indicates that decreasing activity increases risk (an activator/agonist target). Based on the slope sign, gene modulators were classified as activators (negative slope) or inhibitors (positive slope).

#### Benchmarking VIDRA performance with simulations

First, to benchmark the VIDRA Bayesian model performance, we simulated reference dose-response relationships and compared linear regression and two Bayesian models: one using standard regression (where all predictor variables are entered simultaneously) and VIDRA Bayesian model, a hierarchical model that integrates multiple sources of variant information while capturing the unique contribution of each predictor set. To resemble real-world conditions, we evaluated VIDRA’s performance under a simulation framework with a limited number of instrumental variables (bootstrapping between 1 and 5 variants). The simulation introduced variability by assigning different standard deviations to *relative protein function* estimates (ranging from 0.1 to 0.5) and regression error terms (ranging from 0.1 to 0.5), mimicking noise and technical variability from different data sources, such as GWAS, rare coding, and ClinVar (**Methods**).

Under these low-data conditions, VIDRA Bayesian model is a reliable approach for dose-response estimation, demonstrating robust performance when integrating diverse variant sources (Spearman correlation between true and estimated slope, R_VIDRA_ = 0.96 and R_linear-reg_ = 0.64; **Fig. 2a**). The hierarchical aggregation of variant sources into a unified dose-response model produced the best correlation performance, with a mean absolute error of 0.257. Furthermore, VIDRA’s performance remained robust across different numbers of variants and noise levels, achieving R^2^ > 0.8 across the tested parameter ranges (**Supplementary Fig. 2**). Notably, only 3.7% of slope directionalities were misestimated, and these errors primarily occurred when the true slope was close to zero, i.e. in the cases where the phenotype showed minimal dependency on the gene.

**Figure 2.**
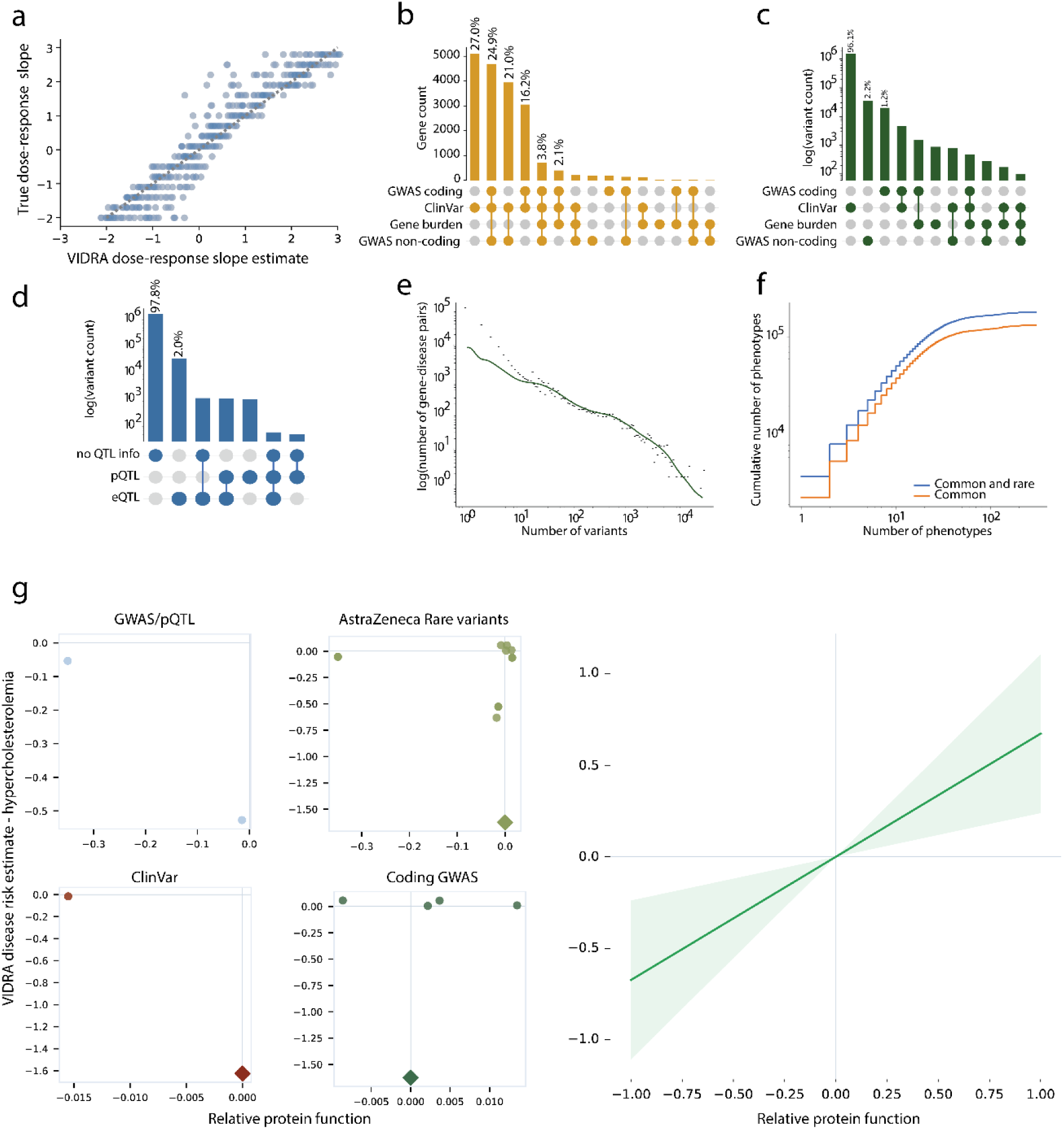
Data overview and benchmarking of VIDRA. **(a)** Performance of slope recovery under simulated low-data conditions (1–5 variants per gene–phenotype pair). Each point represents a simulation; the dotted line indicates perfect recovery (x = y). The hierarchical VIDRA model (dark blue) outperformed both a non-stratified Bayesian model (light blue) and linear regression (grey), achieving Spearman correlations of 0.96, 0.78, and 0.64, respectively. **(b)** UpSet plot showing gene overlap across evidence sources. **(c)** UpSet plot showing variant overlap across evidence sources included in VIDRA. **(d)** UpSet plot of variants with molecular QTL effects on protein abundance or activity across sources. **(e)** Distribution of variant support per gene–phenotype pair. The x-axis shows the number of variants per pair (log scale) and the y-axis shows the number of gene–phenotype pairs (log scale). **(f)** Cumulative number of phenotypes identified by linear and hierarchical models across the same set of genes, demonstrating increased phenotype coverage with hierarchical integration of common and rare variants. **(g)** Example gene–phenotype relationship for *PCSK9* and hypercholesterolaemia. The left panels show variant-level evidence from individual sources (eQTL, pQTL, coding GWAS and ClinVar), with diamonds denoting gene-level burden tests. The right panel shows the integrated VIDRA dose-response estimate across all evidence sources, with the posterior mean slope (green) and 95% credible interval (shaded).

These results establish VIDRA’s Bayesian hierarchical framework as an accurate and reliable method for dose-response estimation while effectively integrating diverse variant sources into the modeling process.

#### Construction of genetic dose-response relationships

To derive genetic dose-response relationships, we used input data of 1,607,115 unique trait-associated variants with estimated *relative protein function* effects in VIDRA regression analyses. We leveraged information for 18,838 genes over 7,681 phenotypes with a median of two disease or clinical phenotypes associated per gene, reflecting the typical sparsity of well-powered genetic associations across the genome. This included 7,437 genes with information from one of the genetic sources (6,997 genes with only rare variants from ClinVar, 400 GWAS only, and 40 with only AZ rare variants). For 11,279 genes, we observed contributions from two or more sources (9,788 ClinVar and GWAS, 259 ClinVar and gene burden, or 35 for GWAS and gene burden). For 1,197 genes, we observed a contribution from all sources (**Fig. 2b**). Out of all the variants assembled for the study, 1,549,806 were rare variants from ClinVar and 63,521 were common variants (**Fig. 2c**). The variant effect estimate on protein (or transcript) abundance was primarily driven by eQTLs (**Fig. 2d**), which is expected given the relatively limited number of available pQTL studies.

In simulated data, the accuracy of inferring allelic series dose-response relationships increased with the number of variants contributing to gene-phenotype pairs. VIDRA achieved a statistical power of alpha 0.8 for slopes ≥ 1 for five or more variants modelled in 29,918 allelic series dose-response relationships (**Supplementary Fig. 2**). In the real data, we observed that the median number of variants describing a gene-phenotype pair was two (**Fig. 2e**). The genes with very high variant numbers were from ClinVar and referred to long proteins (e.g. *TTN*) and germline cancer variants (e.g. *BRCA2*).

In total, VIDRA generated 148,350 dose-response-like gene-phenotype relationships across 7,681 phenotypes. Of these, an estimated 81,163 dose-response relationships (spanning 4,068 phenotypes and 13,305 genes) had posterior probability greater than >0.7 for ClinVar only associations and >0.55 for the multi-sources associations (**Fig. 1; Methods; Supplementary Table 1**). Constraint metric distributions or gene copy number (e.g. LOEUF^36^ or pHaplo^37^) for these genes are not skewed, suggesting the VIDRA score can be informative across multiple genetic constraints (**Supplementary Fig. 3**). Moreover, slope estimates from the VIDRA Bayesian model are independent from the number of variants that the model used to estimate its effect (**Supplementary Fig. 4**). Incorporating rare variant information into the framework led to a 51% increase in the number of unique gene-phenotype associations as compared to using only the common variant information, underscoring the importance of rare variants in this type of analysis (**Fig. 2f)**. Finally, when benchmarking VIDRA’s performance, we observed that it successfully identified the well-established dose-response relationship between *PCSK9* and hypercholesterolemia, accurately capturing the biological link between this gene and the disease. The *PCSK9* dose-response model incorporated eight rare and four common variants (**Fig. 2g**), yielding a VIDRA slope of β=0.67, indicating that reduced *PCSK9* gene activity confers disease protection, a finding consistent with clinical trial outcomes for Alirocumab and Evolocumab, human monoclonal antibodies that target and inhibit PCSK9 protein.^38,39^

### Prioritising novel drug targets using the dose-response relationship

Using the estimated dose-response relationship between genes and phenotypes, we applied a machine learning approach to derive a VIDRA Therapeutic Potential Score to rank drug targets as clinical candidates based on the VIDRA slopes. The VIDRA Therapeutic Potential Score is a classifier trained on features derived from the VIDRA Bayesian model, including the slope estimate, the number of variants supporting it, variant source, and slope posterior confidence, to predict the likelihood that modulating a gene will lead to a successful therapeutic outcome (**Methods**). The distribution of VIDRA Therapeutic Potential Scores emerging from this approach provides insights into how strongly a gene is linked to disease and its potential as a drug target. To ensure its reliability, we trained the VIDRA Therapeutic Potential Score using 2,051 clinically approved drugs with 945 known molecular targets (**Fig. 3a; Methods**). VIDRA Therapeutic Potential Scores range from 0 to 1, scores below 0.6 indicate a weak or inconclusive dose-response relationship that does not support drug development. While genes in this range may still be biologically relevant, they lack strong resemblance to therapeutic targets based on currently developed drugs. Scores between 0.6 and 0.7 represent moderate evidence, identifying potential emerging targets that warrant further investigation. At a Therapeutic Potential score ≥ 0.6, VIDRA prioritises 6,384 genes that currently are not approved drug targets, while a more stringent Therapeutic potential score ≥ 0.7 selects 1,637 genes. These genes have VIDRA slope profiles closer to those of approved drug targets, consistent with their being plausible therapeutic targets. In fact, increasing the threshold stringency on VIDRA Therapeutic Potential Score enriches successful drug targets, with more than 10-fold and 24-fold enrichment in approved drugs, for scores above 0.6 and 0.7 respectively (Fisher’s exact test, OR = 10.73, p-value = 3.85 x 10^-104^; OR = 24.86, p-value = 2.78 x 10^-49^; **Fig. 3b**). This large enrichment could be explained by the inclusion of genes that are linked to rare inherited disorders, which have shown larger success rate also in previous studies.^5^ This enrichment was independently validated using a complementary, more conservative framework that matches gene–indication pairs directly to clinical trial outcomes, with drug–indication links propagated through the EFO/MONDO disease ontology; this analysis confirmed significant, threshold-dependent enrichment of approved Phase IV targets (**Supplementary Fig. 5**). Finally, we compared VIDRA performance against the genetic priority score (GPS) method^2^ that prioritizes drug targets by integrating eight phenotype-specific features derived from clinical, coding, and genome-wide association genetic evidence. We found that VIDRA outperformed GPS (**Supplementary Fig. 6**), achieving up to a 24-fold enrichment for prioritising genes with known drug indications, compared to an 11-fold enrichment by GPS workflow.^2^

**Figure 3.**
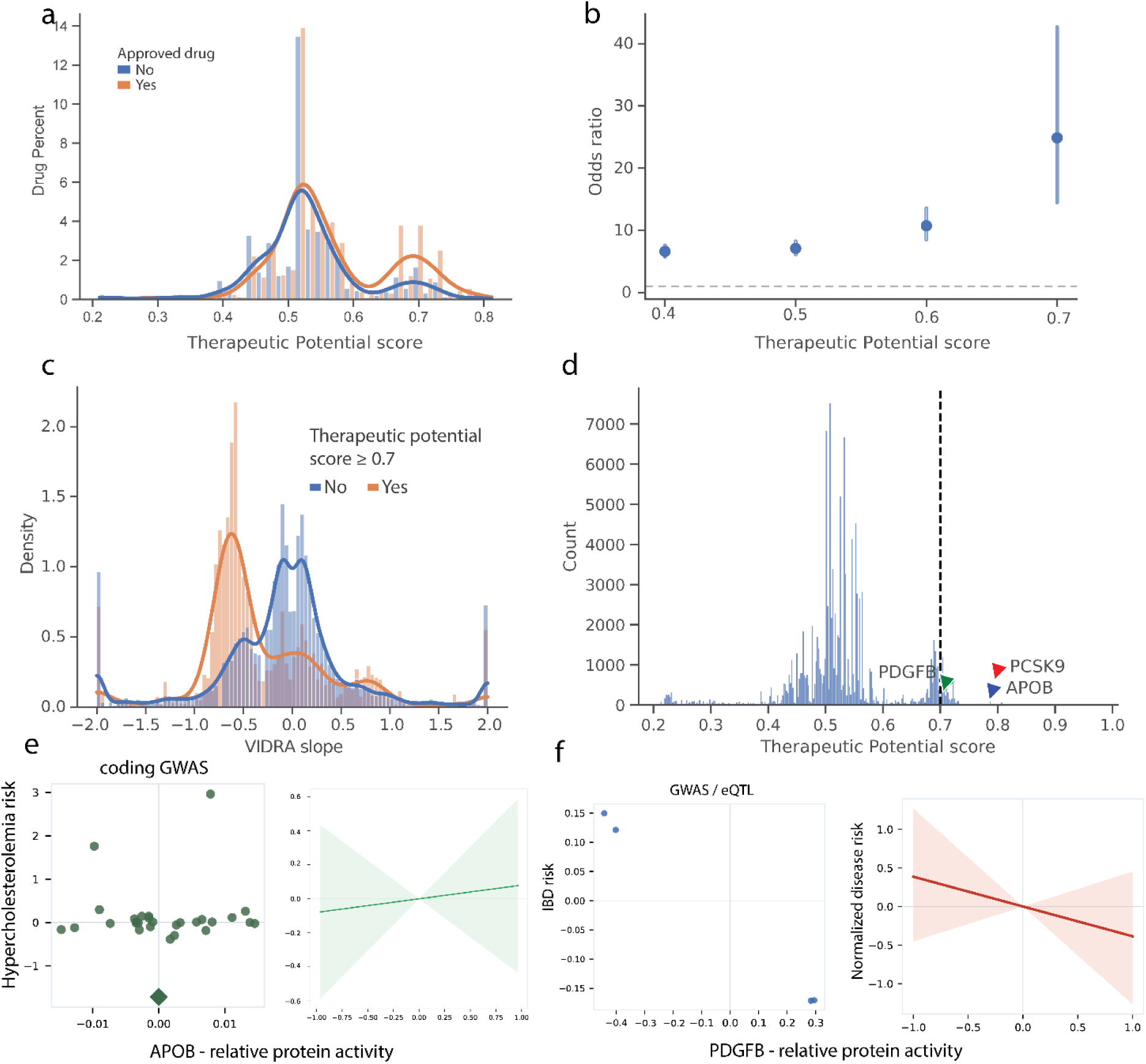
VIDRA Therapeutic Potential Score prioritises clinically successful drug targets. **(a)** Distribution of VIDRA Therapeutic Potential Scores for drug targets stratified by clinical trial outcome (approved versus not approved). **(b)** Odds ratio of drug approval across increasing Therapeutic Potential Score bins, showing higher likelihood of clinical success for highly prioritised targets. **(c)** Distribution of VIDRA slopes stratified by Therapeutic Potential Score (>0.7 versus ≤0.7). Highly prioritised gene–phenotype pairs are enriched for larger-magnitude slopes, indicating stronger gene–phenotype dependency. **(d)** Distribution of Therapeutic Potential Scores across all gene–phenotype pairs. The dashed line indicates the prioritisation threshold of 0.7. **(e)** Example association between *APOB* and hypercholesterolaemia. Left, variant-level evidence from individual sources, with diamonds denoting gene-level burden tests. Right, integrated VIDRA dose-response estimate showing the posterior mean slope (green) and 95% credible interval (shaded). **(f)** Example association between *PDGFB* and inflammatory bowel disease, displayed as in (e). The posterior mean slope is shown in red with the corresponding 95% credible interval.

VIDRA captures both positive and negative slopes across all evidence sources, inferring directionality from the data, rather than imposing it on the model. Generally, we observed a similar distribution of variant sources across the VIDRA slope spectrum, with the exception of ClinVar and ClinVar + GWAS coding that were enriched in the negative slopes (**Methods** and **Supplementary Fig. 7)**. Gene–trait pairs with a high Therapeutic Potential Score (≥ 0.7) show a bimodal distribution of VIDRA slopes, with peaks at approximately −0.6 and +0.8, whereas pairs with a low score show a unimodal distribution centred near zero (**Fig. 3c**). The modest asymmetry towards negative slopes among high-scoring pairs mirrors the predominance of inhibitory mechanisms among approved drugs, reflecting the composition of the training data rather than a biological prior, and underscoring the untapped therapeutic opportunity in the activator space. This systematic shift towards larger-magnitude VIDRA slope magnitudes, in both directions, among highly prioritised gene–trait pairs reflects stronger causal dependency between gene function and phenotype, consistent with a more pronounced phenotypic consequence of gene perturbation. Although modest, we observed that VIDRA slope magnitude correlated positively with Therapeutic Potential Score within each trait with at least two associated genes (Spearman R = 0.58, 95%CI: 0.45-0.9; **Supplementary Fig. 8**). These observations support the interpretation that the VIDRA Therapeutic Potential Score preferentially identifies genes with pronounced functional dependency on phenotype, a property associated with a greater likelihood of therapeutic success.

Therefore, focusing drug development on genes that significantly affect phenotypes may translate to a higher likelihood of successful drug development.^40^ Among the high-scoring genes, which are drug targets, we identified *TYK2* (VIDRA Therapeutic Potential Score = 0.61) and *PCSK9* (VIDRA Therapeutic Potential Score = 0.80), two hallmark examples of drug targets with strong genetic evidence, supported by common and rare variant associations, respectively.^41–44^ Notably, the few high-scoring targets (VIDRA Therapeutic Potential Score > 0.7) that have not yet received regulatory approval are currently in early clinical trial development, 9 in phase 1 and 14 in phase 2, suggesting they still have the potential to succeed (**Supplementary Fig. 9**).

Expanding VIDRA therapeutic scoring approach, beyond the curated set of gene-phenotypes with known drugs and clinical candidates, across the full VIDRA framework dataset, we identified 1,637 genes for 1,287 possible indications with a VIDRA Therapeutic Potential Score ≥ 0.7 which were not previously reported as approved drug targets (**Fig. 3d; Supplementary Table 2**). For instance, our approach highlighted *APOB*, a less commonly discussed but genetically well-supported target. *APOB* (VIDRA Therapeutic Potential Score = 0.81), similar to *PCSK9*, is the target of approved cholesterol-lowering therapies used to treat hypercholesterolemia. Population-scale association studies have linked *APOB* variants to hypercholesterolemia, with familial forms caused by rare variants.^45,46^ Common variants, such as rs1367117, are associated with increased APOB expression (i.e., gain-of-function), leading to an elevated risk of hypercholesterolemia.^47,48^ Similarly, rare variants such as Arg3500Gln result in higher apolipoprotein B blood levels, increasing the risk of hypercholesterolemia and ischemic heart disease.^45,49^ The APOB-targeting drug Mipomersen functions through an mRNA-binding mechanism, reducing protein expression and lowering cholesterol levels, a therapeutic direction of modulation also captured by the VIDRA dose-response model (**Fig. 3e**).^50,51^

Beyond well-established targets, VIDRA also identifies potential novel drug targets, such as *PDGFB* VIDRA Therapeutic Potential Score = 0.61). Variants in *PDGFB* have been identified in several eQTL studies, showing both positive and negative effects on gene expression,^52–54^ with colocalization analyses linking them to variants that either protect against or increase the risk of inflammatory bowel disease (IBD) (**Fig. 3f**).^55,56^ Genetic studies have further associated *PDGFB* with key IBD-related processes, including angiogenesis and macrophage-mediated inflammation, contributing to IBD pathogenesis.^57,58^ Notably, *PDGFB* is already a target of drugs in clinical development, with some reaching Phase 3 trials, suggesting potential opportunities for drug repurposing in IBD.^59,60^

### VIDRA Bayesian model slope estimates inform drug therapeutic direction of modulation

The directionality of VIDRA slopes provide insights into potential drug therapeutic direction of modulation. A negative slope suggests that reduced protein function increases disease risk, indicating that therapeutic strategies should aim to increase or stabilise protein activity. Conversely, a positive slope implies that reduced protein function confers disease protection, serving as a proxy for an inhibiting therapeutic direction of modulation. For non-coding variants with eQTL or pQTL colocalisation, QTL effect sizes capture both directions of gene expression or protein level change, enabling VIDRA to integrate gain- and loss-of-function effects simultaneously. For coding variants annotated by *in silico* pathogenicity tools, the exposure axis is currently restricted to the loss-of-function spectrum, as available predictors are predominantly trained on loss-of-function examples and have limited capacity to classify gain-of-function variants reliably; gain-of-function effects for this class are captured indirectly through the sign of the resulting slope.

To validate our approach, we benchmarked VIDRA’s ability to predict the direction of the modulation against a set of 551 approved drugs with known targets and therapeutic direction of modulation. We observed high sensitivity and specificity in VIDRA slope-derived classification of therapeutic direction of modulation (area under the ROC curve, AUC, of 0.70 for agonists and 0.72 for antagonists across all therapy areas; **Fig. 4a**), outperforming the GPS (**Supplementary Fig. 6c)**. The AUC improves at more stringent VIDRA slope thresholds, indicating that larger-magnitude slopes more reliably predict the direction of target modulation. However, the perfect classification at the most stringent threshold (AUC = 1.00) should be interpreted cautiously, as only 48 gene–phenotype pairs were retained and the result may partly reflect limited sample size (**Supplementary Fig. 10**). Finally, to assess the significance of these AUC estimates, we performed a permutation test by randomly reshuffling the therapeutic direction of modulation labels 10,000 times, yielding a p-value of 10^-4^ for both inhibitors and activators (**Supplementary Fig. 6b**). These results reinforce the confidence in VIDRA’s ability to correctly infer drug therapeutic direction of modulation.

**Figure 4.**
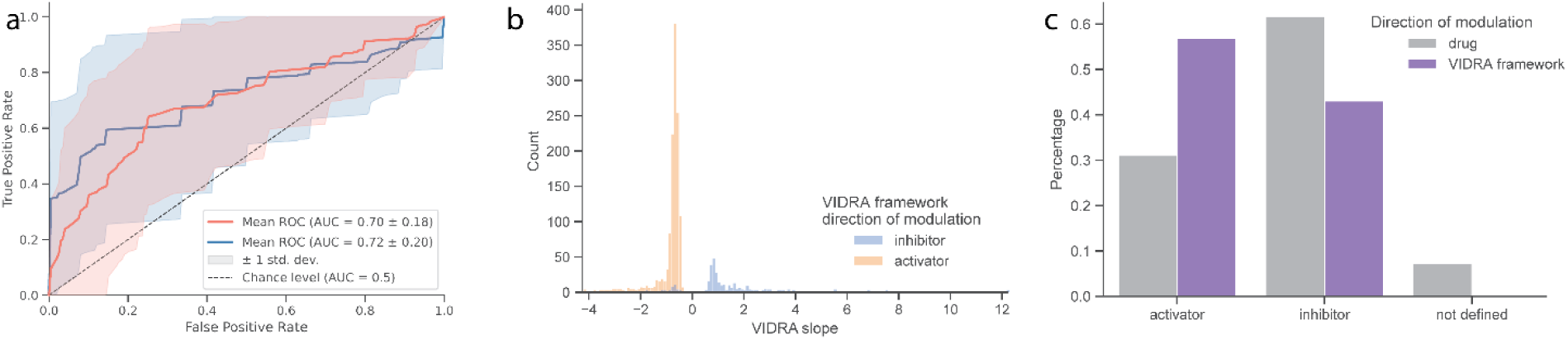
VIDRA slopes infer therapeutic direction of modulation. **(a)** Performance of VIDRA slope sign for predicting therapeutic direction of modulation. Receiver operating characteristic (ROC) curves are shown separately for inhibitors (blue) and activators (orange) across therapy areas. Shaded bands indicate ±1 standard deviation; the dashed diagonal indicates chance performance. **(b)** Distribution of VIDRA slopes stratified by inferred therapeutic direction of modulation (activator versus inhibitor), illustrating how slope sign determines the predicted therapeutic direction of modulation. **(c)** Comparison of the proportions of activators, inhibitors and undefined mechanisms among approved drugs using external annotations (grey) and VIDRA-derived predictions (purple). Undefined mechanisms correspond to approved drugs whose therapeutic direction of modulation could not be harmonised into activator or inhibitor categories.

Expanding this analysis to all gene-phenotype pairs in our dataset, we assigned a plausible therapeutic direction of modulation to the 1,860 target genes identified using a VIDRA Therapeutic Potential Score threshold of ≥ 0.7 (**Fig. 4b**). The majority (57%) of these predictions suggest that therapeutic intervention would require an agonistic action, aligning with genetic observations that loss-of-function variants are the most commonly implicated pathogenic mechanism (**Fig. 4c**).^61,62^ Interestingly, this agonist/antagonist ratio is the opposite of the current distribution of approved therapeutic direction of modulation, where 62% of approved drugs function as antagonists (**Fig. 4c**).^63,64^ This discrepancy reflects a well-known trend in drug development: inhibiting a target is generally more feasible than enhancing its activity.

### VIDRA dose-response identifies disease biomarkers, adverse events and informs drug dosing

Most genetic approaches in drug discovery primarily focus on target prioritisation. However, genetic data can also address other critical aspects of drug development, such as identifying disease biomarkers or identifying pleiotropic effects, which in turn, can inform on the risks of modulating a target and the development of other diseases. We hypothesised that VIDRA could extend beyond target selection to inform these aspects by establishing relationships between genes, biomarker measurements, and disease occurrence, thereby providing insights into biological mechanisms and adverse effects.

First, to assess how VIDRA slope estimates could aid biological interpretation for drug discovery, we examined correlations between established biomarker measurement-disease relationships and their respective VIDRA slopes. We manually curated a list of 572 known biomarker-disease pairs based on established knowledge. This accounted for 962 genes associated with these biomarker-disease pairs, with 59% having an effect on two or more biomarker-disease pairs (**Supplementary Table 3**). The pairs included, for example, relationships such as cholesterol levels and cardiovascular disease, blood glucose levels and type 2 diabetes, platelet aggregation and stroke, and others, capturing a broad representation of biological systems and phenotypes. We hypothesised that when a biomarker has positive correlation with disease risk, VIDRA framework should capture a positive relationship between their VIDRA slopes (e.g. cholesterol levels and cardiovascular disease). Conversely, when a biomarker measurement is inversely correlated with disease risk (e.g. platelet count and bleeding risk), we expect to observe opposite slope directionality. Our analysis revealed a positive correlation in cases where both the biomarker and disease share the same directionality (both positive: β=0.33, p_perm_=1×10^-4^; or both negative: β=0.41, p_perm_=1×10^-4^; **Fig. 5a** and **Supplementary Fig. 11**) and a significant negative correlation when they exhibit opposite effects (increased disease risk and reduced biomarker: β=-0.31,p_perm_=1×10^-4^; reduced disease risk and increased biomarker: β=-0.14, p_perm_=1×10^-4^; **Fig. 5b** and **Supplementary Fig. 11**). To assess statistical significance, we randomly permuted 1,000x labels between biomarker-disease pair slopes to generate a null distribution (**Supplementary Fig. 11**). VIDRA accurately captures well established correlations such as the negative correlation between HDL cholesterol levels and coronary atherosclerosis^65,66^ or the positive correlation between waist circumference and obesity (**Fig. 5a** and **Fig. 5b**).^67,68^

**Figure 5.**
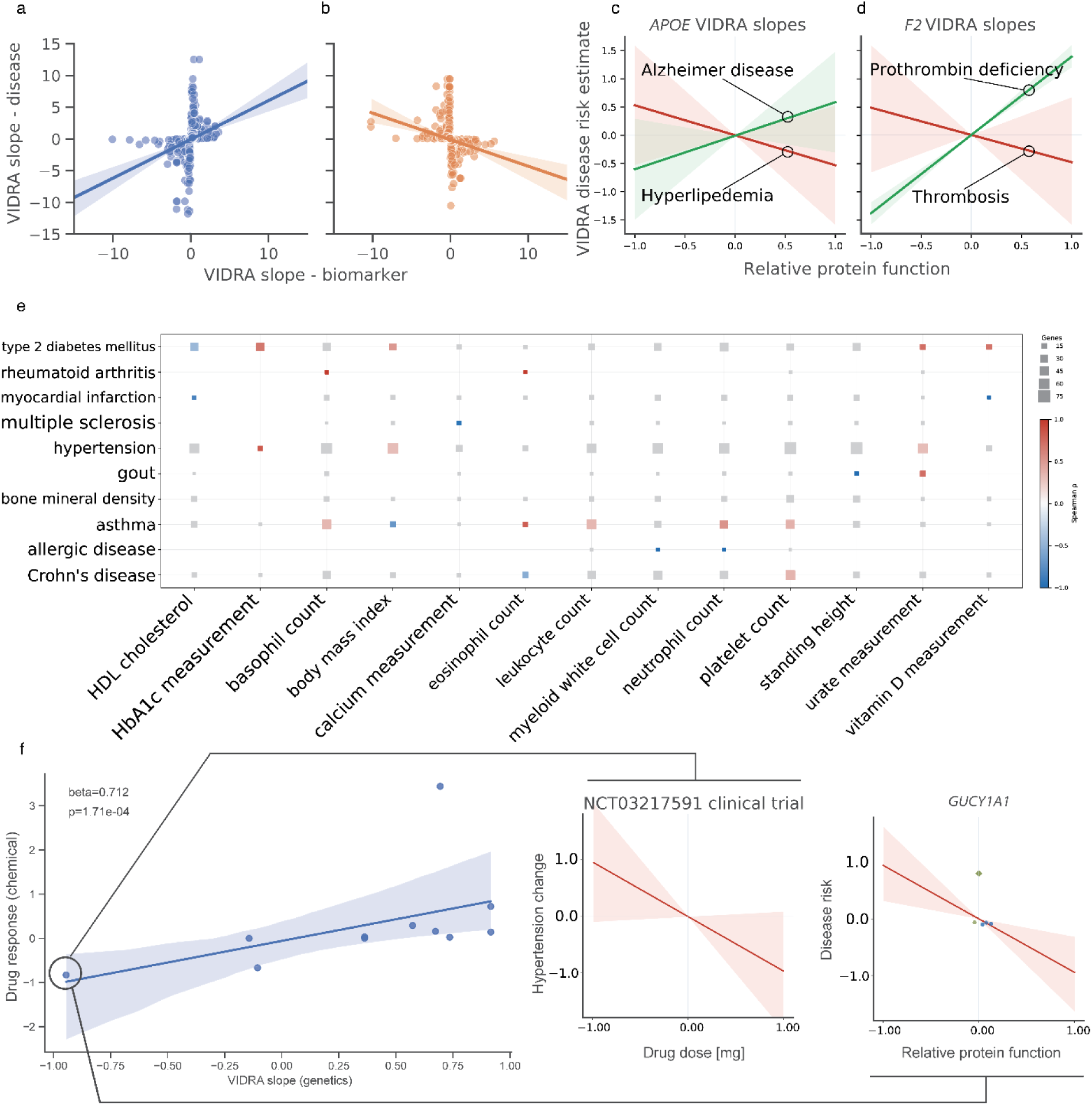
VIDRA slope estimates capture biological relevance between phenotypes and biomarkers, identify windows of target modulation, and predict potential side effects. **(a)** Correlation between VIDRA slope estimates for intermediate phenotypes (x-axis) and disease phenotypes (y-axis) from the curated set in Supplementary Table 3, showing gene–phenotype pairs with positive slope concordance. **(b)** as in **(a)** but showing negative slope concordance. **(c)** Example of therapeutic trade-offs, where modulating a single gene may improve one phenotype while increasing the risk of an adverse event, illustrated with APOE showing opposing VIDRA slopes for hyperlipoproteinemia and Alzheimer’s disease; the x-axis represents relative protein function and the y-axis represents disease risk; shaded areas indicate 95% credible intervals. **(d)** Example of adverse effects from excessive target modulation illustrated with F2, showing slopes for prothrombin deficiency and thrombosis, in the same format as **(c)**. **(e)** Heatmap showing Spearman correlation coefficients between VIDRA slopes for intermediate biomarker phenotypes (x-axis) and a set of human disease phenotypes (y-axis); tile colour indicates direction of correlation (red = positive, blue = negative) and tile size is proportional to the number of genes supporting the association; association not-statistically significant are shown in gray. f) Regression analysis comparing VIDRA slopes (x-axis) with drug response across different drug doses retrieved from clinical trials (y-axis). The shaded blue area represents the 95% confidence interval. The circled data point highlights an example of *GUCY1A1* and its ligand Praliciguat, with an inset showing the underlying distribution of observations contributing to positive correlation.

As gene-phenotype association data continue to expand, it is becoming increasingly feasible to systematically identify genetically supported disease biomarkers. Among these, circulating blood and metabolic measurements are widely used for the diagnosis, prognosis, and monitoring of a broad range of diseases, and many are highly polygenic with abundant genetic association data. To explore whether the VIDRA framework can leverage this information systematically, we extended our analysis to assess relationships between 13 intermediate phenotypes, including blood cell counts, glycaemic markers, lipids, anthropometric measurements, and circulating minerals, and 10 disease phenotypes sharing gene associations with these traits (**Fig. 5e; Supplementary Table 4**). We successfully recovered a broad (n=25, 19%) set of significant (Spearman correlation coefficient p-value < 0.05) established biomarker-disease relationships. We observed numerous positive control relationships, such as the positive correlation between HbA1c and type 2 diabetes (Spearman ρ = 0.85, p = 1.3×10^−8^, n = 28 shared genes),^69^ urate measurement and gout (ρ = 0.72),^70^ or eosinophil count and asthma (ρ = 0.69, p = 0.040, n = 9).^71^ VIDRA framework also captured negative known correlations, such as HDL cholesterol and myocardial infarction (ρ = −0.89) and with type 2 diabetes (ρ = −0.45).^72^ Beyond these known associations, the analysis identified several biologically coherent relationships that extend the originally reported findings. For example, calcium measurement showed a positive correlation with diastolic blood pressure (ρ = 0.50), in line with the role of calcium-handling genes as shared GWAS loci for vascular reactivity and blood pressure regulation.^73^ HbA1c also correlated with hypertension (ρ = 0.67), consistent with shared cardiometabolic biology.^74^ BMI showed an unexpected negative correlation with asthma (ρ = −0.77), contrary to MR evidence supporting a causal effect of higher BMI on asthma risk^75,76^. This discordance may reflect heterogeneity in asthma GWAS definitions, non-linear BMI–asthma relationships, or the small number of shared genes captured by VIDRA. It may also indicate that the BMI-associated genes represented here are enriched for metabolic rather than inflammatory or mechanical pathways implicated in asthma. Together, these results show that VIDRA recovers established biomarker–disease relationships, identifies genetically supported connections across phenotypes, and highlights cases where pathway specificity may limit generalisability.

Extending the logic of identifying biomarkers across broad trait groups, we hypothesised that opposite slope directionalities for two or more phenotypes can provide insights into potential adverse events. That is where therapeutically modulating a target to improve one phenotype may simultaneously increase the risk of an unintended outcome due to the target’s distinct roles across multiple traits. An example of such risks is *APOE*, which encodes a protein involved in lipid transport and is genetically linked to Alzheimer’s disease risk.^77,78^ However, reducing ApoE protein levels is also associated with an increased risk of cardiovascular disease due to elevated circulating cholesterol and triglycerides.^79^ This opposing biology is reflected in the *APOE* VIDRA slope for hyperlipoproteinemia (VIDRA slope β = −0.53) and Alzheimer’s disease (VIDRA slope β = 0.35) demonstrate opposite effects (**Fig. 5c**).

In addition to direction of target modulation, another critical aspect in successful therapeutic intervention is the magnitude of target modulation, while moderate target modulation might be beneficial for treating the intended condition, excessive perturbation could increase the likelihood of adverse effects. A pattern that can be observed in F2, a key protein in the coagulation cascade, which is associated with thrombosis and bleeding risk.^80,81^ The two phenotypes result from opposing functional states of F2: excessive activity increases the risk of thrombosis, while insufficient activity leads to bleeding due to inefficient coagulation. This bidirectional effect is captured in the opposing VIDRA slopes for *F2* gene, for thrombosis (VIDRA slope β=0.02) and prothrombin deficiency (VIDRA slope β=-0.43; **Fig. 5d**). Notably, this biology aligns with clinical observations, as thrombosis treatments like Lepirudin and Bivalirudin carry bleeding risks as side effects.

Finally, understanding the dose-response relationship between gene function and phenotypic outcome is a crucial aspect of drug development for optimizing drug dosing and therapeutic intervention windows. VIDRA slopes capture the quantitative effect of genetic variation on phenotype, which can be leveraged to infer the potential consequences of the different levels of modulating a drug target, i.e. as genetic variants mimic pharmacological perturbations, we hypothesised that VIDRA slopes can provide a genetic proxy for drug titration effects. To explore this relationship, we manually curated clinical trial reports for a subset of 11 drugs with well-characterized molecular targets (**Supplementary Table 5**) and inferred their dose-response relationships at different drug doses. Such analysis suffers from sparsity, nevertheless it revealed a positive correlation (robust linear model β=0.71, p-value=1.17×10^-04^) between the therapeutic effect of a chemical drug as a function of its dose and the corresponding dose-response derived from VIDRA slope estimates (**Fig. 5f**). This finding suggests that genetic evidence, particularly the strength of the dose-response slope, may serve as a proxy for pharmacological sensitivity of a target, helping to estimate the effective drug doses, minimizing the risk of underdosing (inefficacy) or overdosing (toxicity). By aligning VIDRA slope estimates with real-world pharmacokinetic and pharmacodynamic data, genetic evidence could enhance early-stage dose selection, reduce trial-and-error dosing strategies, and improve the design of safer, more effective treatments.

## Discussion

In this study, we used naturally occurring variants identified in common and rare disease studies to model the dose-response relationship between protein activity and the risk of developing diseases. VIDRA framework demonstrated that slope estimates from its Bayesian model can inform several steps of the drug development workflow, from target prioritisation through prediction of adverse effects of target modulation to drug dosing.

Our analysis revealed that genes with steeper dose-response slopes are more likely to be successful drug targets. This suggests that when a phenotype is strongly dependent on gene function, pharmacological modulation of that function is more likely to yield clinically meaningful outcomes. Slope estimates from VIDRA are consistent with previous observations that drug targets enriched in ClinVar or gene burden data, both of which enrich for loss-of-function variants with large biological effects, are more likely to succeed in the clinic.^2^ Incorporating rare variant information into the VIDRA framework resulted in a 51% increase in gene-phenotype associations compared to using only common variants, highlighting the critical contribution of rare variants to this type of analysis. Importantly, our results are in line with previous studies showing comparable drug success rates across a spectrum of variant frequencies and effect sizes^3^, as VIDRA slopes reflect a functional dependency rather than the absolute genetic effect of variants. Indeed, even variants with modest effect sizes can imply a strong dose-response relationship if they align consistently across multiple sources of genetic evidence. Moreover, the bimodal distribution of VIDRA Therapeutic Potential Scores, with a subset of successful drug targets scoring just below our 0.7 threshold, highlights the potential for further refinement. As genetic association data continue to grow and functional insights from variant perturbation experiments, such as saturation genome editing, expand our understanding of how variants impact protein function, incorporating these additional layers of evidence will enhance the model’s resolution and significantly improve its predictive utility.

To date, the use of human genetics in drug discovery has largely focused on identifying and prioritizing novel drug targets, particularly those supported by genome-wide association studies and rare variant analyses.^35^ However, in this study, we demonstrate that genetic data, when structured as allelic series, can inform multiple additional aspects of the drug development pipeline. For example, our dose-response modelling approach provides early estimates of a drug’s likely direction of modulation, which could be used to prioritise experimental assays and streamline target validation workflows. Having prior insight into whether a target is more likely to benefit from activation or inhibition allows for more focused experimental design and potentially faster progression to proof-of-concept studies.

Furthermore, our analysis further reinforces the mismatch between the proportion of targets that genetically suggest a need for agonist drugs and the current composition of approved therapeutics, which are predominantly inhibitors. This is explained by the fact that it is much easier to achieve an inhibitory or suppressive modulation of a target but it emphasises that there is substantial untapped opportunity to expand the pool of agonist drugs. Developing robust and scalable methods for activating targets, both chemically and biologically, remains an area for innovation.

VIDRA’s dose-response estimates also capture biologically meaningful relationships that extend beyond simple target prioritization. First, we show that these estimates align with intermediate phenotype-disease correlations, a feature that may be used to identify novel disease biomarkers or monitor drug efficacy. Second, the relationship between VIDRA slopes and clinical drug dosing supports the idea that genetic dose-response modelling can inform therapeutic windows and aid dose selection in early-phase trials. Lastly, by detecting opposite directionalities of effect across multiple phenotypes for the same gene, VIDRA can anticipate on-target adverse effects, highlighting risks of therapeutic trade-offs or dosing-related toxicity.

Despite these advances, we acknowledge several limitations. The analysis depends on the variants and phenotypes catalogued in the Open Targets Platform, which may reflect ascertainment biases. For example, some associations may be missing due to underpowered studies, limited phenotype coverage, or may simply not be deposited in the open source resources that Open Targets relies on. Additionally, many filtering steps in the Open Targets genetics analysis pipeline rely on predefined thresholds, which could exclude relevant but borderline signals. The inclusion of rare variants increased the number of allelic series estimates by 34.5%, but the use of *in silico* predictions to annotate these variants introduces the potential for misclassification, which could propagate into the dose-response models. Moreover, gene burden tests are currently limited by cohort size, variant annotation accuracy, and phenotyping resolution, which can lead to inconsistent signal strength or false negatives, particularly for genes with modest or context-dependent effects. Finally, the available data is based on mainly European cohorts, therefore the relevance across ancestries remains to be assessed.

A key limitation of the current approach lies in how variant effects on gene function are defined and interpreted. In VIDRA, we assume a linear relationship between inferred gene function and phenotype, a pragmatic choice driven by data sparsity, which can describe only a small linear portion of the sigmoid. With a median of two variants per gene–phenotype pair, more complex non-linear models do not converge reliably and risk overfitting. Classic dose–response relationships are often described by Hill functions, which model sigmoid dynamics including cooperativity, thresholds, and saturation. These features may be particularly relevant in systems where small perturbations in gene function have disproportionate phenotypic consequences or where effects plateau beyond a certain activity level. As variant-to-function maps from resources such as saturation genome editing continue to expand, future iterations of the model could incorporate such non-linear frameworks to improve biological interpretability. A related constraint concerns the directionality of variant effect estimates for rare coding variants. Current in silico predictors are trained predominantly on loss-of-function (LoF) examples and retain limited capacity to classify gain-of-function (GoF) variants reliably; we therefore restrict variant effect scores to the 0–1 LoF spectrum. Within VIDRA, GoF effects for rare-coding variants are captured indirectly: genes where LoF variants confer disease protection yield positive slopes, implying that increased gene activity drives disease risk and that inhibition is the appropriate therapeutic direction. While biologically principled, this means that genes driven primarily by direct GoF coding variants may be underrepresented in the current framework. Expanding variant effect annotations to encompass activating mutations, through resources such as saturation genome editing, deep mutational scanning, and emerging gain-of-function classifiers, will be essential for removing this asymmetry and improving VIDRA’s coverage of genes whose disease biology is driven primarily by direct gain-of-function coding variants.

Furthermore, for common variants, we rely primarily on steady-state eQTL data, which represent gene expression under baseline conditions. However, it is well established that many regulatory effects are context-dependent, manifesting only under specific stimuli, developmental stages, or environmental exposures.^19,52,54^ Moreover, eQTLs often show limited correlation with protein levels, and thus can only serve as a proxy for protein activity. This limitation is further compounded by the fact that our eQTL colocalisation analyses were restricted to blood and immune cell types, where data are most abundant. While these tissues are relevant to many diseases, expanding the model to incorporate QTLs from other tissues will be crucial for improving resolution across a broader range of phenotypes. Furthermore, variants affecting splicing can have profound consequences on protein isoform diversity and function. Integrating splice-QTLs and isoform-specific expression data will enhance our ability to connect genetic variation to functional protein outcomes.

In the context of drug development, temporal dynamics are also critical. Real-world pharmacology involves time-dependent exposure and clearance, whereas genetic models such as VIDRA are based on lifelong germline variant effects. Incorporating somatic variation, which can model tissue-specific or time-limited changes in gene function, may offer a path forward for refining these models and bringing them closer to clinical pharmacokinetics and pharmacodynamics. In parallel, there is a growing need to improve the functional annotation of all classes of coding variants, including missense and synonymous. As the field advances, resources like saturation genome editing experiments will provide high-resolution, quantitative maps of variant effects on gene function. Incorporating such data will improve the classification of variants of uncertain significance and refine predictions of dose-response relationships. Ultimately, to capture the full biological impact of genetic variants, effects on modulating cellular functions should also be captured. However, resources linking variant-level perturbations to cell-level outcomes remain scarce, and addressing this gap will require community-scale efforts and the development of systematic, high-throughput functional genomics platforms.^82–84^ As these resources grow, they will enable increasingly accurate, context-specific models that bridge the gap from variant to function and from gene to therapeutic intervention.

VIDRA’s biomarker analysis is inherently constrained to biomarker-disease relationships with a detectable germline genetic component. Biomarkers predominantly reflecting acute physiological responses or environmental exposures may lack sufficient heritable genetic architecture to be captured by this framework. The curated biomarker-disease pairs used in this analysis were selected to represent traits with established genetic associations, spanning multiple biological systems, including metabolic, cardiovascular, haematological, and immunological domains; pairs without meaningful GWAS or rare variant evidence were excluded by design.

Finally, phenotype harmonization also posed challenges. Using Experimental Factor Ontology (EFO) codes to approximate clinical phenotypes introduced semantic noise and redundancy, particularly when biologically similar traits were split into distinct terms. This duplication can reduce power and obscure shared biological signals. Moreover, refined curation of EFO would also improve the adverse events definition. For example, a gene with a negative slope for platelet count will also show a positive slope for thrombocytosis, not due to side effects, but because of true biology. Currently, these connections are not captured in the most common ontologies. Improved ontology curation, especially efforts that unify related traits and better capture relationships between common and rare disease phenotypes, would significantly enhance analyses like this. Finally, studies of quantitative traits often apply transformation to make traits more normally distributed, which can alter the scale and make interpretation of the effect sizes harder to compare across studies or to interpret biologically.

In summary, we present a framework for integrating genetic allelic series into multiple stages of drug development, from target prioritization to therapeutic direction of modulation prediction, biomarker discovery, dose guidance, and safety risk assessment. While the current version of VIDRA benefits from harmonised data and robust statistical modelling, it also stands to gain from the incorporation of additional functional variant classification, richer phenotype definitions, and more diverse genomic resources. The growth of large-scale functional genomics projects and the expansion of diverse biobanks across ancestries and phenotypes will enable future iterations of this framework to become more predictive, more generalizable, and more useful for translating genetic insights into therapeutic advances.

## Methods

### Data availability statement

1. GitHub Repository: https://github.com/TrynkaLab/VIDRA
2. Open Targets Genetics: https://ftp.ebi.ac.uk/pub/databases/opentargets/genetics/22.09/
3. Open Targets Platform: https://ftp.ebi.ac.uk/pub/databases/opentargets/platform/24.03/
4. Input data for the model, including variants and their annotations, burden tests and drug information have also been made available at: https://www.ebi.ac.uk/biostudies/studies/S-BSST3098 under the CCO license. Please note that the same data are also available and maintained in the Open Targets platform.

### Chemical compound and clinical trial data

Information on the chemical compounds, molecular targets and clinical phase was collected by ChEMBL and obtained via the Open Targets Platform using the BigQuery dataset ‘open-targets-prod.platform.molecule’, downloaded on 31/10/2023.^7^ We defined drugs as approved when they achieved clinical trials in phase 4 (1,484 drugs for 1,788 indications in 453 genes), while we considered not-approved drugs with clinical trials in phase 3 or below (1,786 compounds for 811 indications in 797 genes). Information on the drug direction of modulation came from the BigQuery dataset ‘open-targets-prod.platform.mechanismOfAction’ downloaded on 31/10/2023. Drugs labelled as RNA inhibitors, negative modulators, negative allosteric modulators, antagonists, antisense inhibitors, blockers, inhibitors, degraders, inverse agonists, allosteric antagonists, and disrupting agents were defined as inhibitors. Drugs labelled as partial agonists, activators, positive allosteric modulators, positive modulators, agonists, sequestering agents or stabilisers were defined as activators. Information on the chemical dose response was obtained via manual curation of the clinical trial results reported in https://clinicaltrials.gov/. The list of curated clinical trials, chemical doses and measured outcomes is reported in **Supplementary Table 5**.

### GWAS and eQTL data

The variant information used in this study was obtained from Open Targets Platform and Genetics, version 24.03 and 22.09 respectively, via the BigQuery dataset open-targets-genetics, downloaded on 20/03/2024.^7^ Common causal variants were identified through fine-mapping when summary statistics were available, or via LD expansion when they were not. GWAS lead variants were retained if both the conditional and nominal p-values were less than 5 x 10^−8^. For molecular QTL lead variants, a more lenient threshold was applied to account for reduced statistical power: both nominal and conditional p-values had to be less than 0.05 divided by the number of variants tested per gene.

Summary statistics were fine-mapped using approximate Bayes factor method and 95% credible sets were defined.^85^ Colocalisation analysis was then performed between overlapping credible sets to identify independent variants sharing a causal signal, with one variant coming from a disease GWAS and the other from a molecular QTL. The VIDRA workflow incorporates only those variants with colocalisation posterior probability (coloc H4) > 0.7.

When molecular QTL data were available, we focused specifically on QTLs derived from blood tissue and immune cell types, given that these represent the largest and most well-powered eQTL datasets. We selected the tissues using the following labels: uberon_0000178, blood, uberon_0001969, treg_naive, treg_memory, th2_memory, th1_memory, th17_memory, th1-17_memory, tfh_memory, nk-cell_naive, monocyte_naive, monocyte_cd16_naive, cd8_t-cell_naive, cd8_t-cell_anti-cd3-cd28, cd4_t-cell_naive, cd4_t-cell_anti-cd3-cd28, b-cell_naive, monocyte_r848, monocyte_pam3csk4, monocyte_lps, monocyte_iav, macrophage_salmonella, macrophage_naive, macrophage_listeria, neutrophil_cd16, t-cell_cd8, t-cell_cd4, neutrophil, monocyte, macrophage_ifng, macrophage_ifng+salmonella, t-cell, monocyte_lps24, monocyte_lps2, monocyte_ifn24, b-cell_cd19, platelet, neutrophil_cd15, monocyte_cd14. In cases where we observe colocalisation across multiple cell types, we selected the cell type with the most significant p-value to retrieve effect size information for downstream modelling.

#### Gene Burden datasets

UK Biobank 450K exome sequencing gene burden summary statistics were obtained directly from the raw AZ PheWAS collapsing model data (binary and quantitative phenotypes), obtained from the AstraZeneca PheWAS Portal (https://www.azphewas.com/)^32^ and accessed through the Open Targets Platform (version 22.04).^7^ We retained burden results from three collapsing model categories: (i) protein-truncating variants with MAF ≤ 0.001 in both the cohort and gnomAD (ptv); (ii) protein-truncating variants with MAF ≤ 0.05 in both the cohort and gnomAD (ptv5pcnt), noting that this more lenient frequency threshold may introduce greater susceptibility to population stratification confounding; and (iii) a combined model aggregating moderately rare PTVs (MAF ≤ 0.001) and rare non-synonymous variants predicted damaging by REVEL score ≥ 0.25 (non-PTV MAF ≤ 0.00025 in cohort, gnomAD global_raw ≤ 0.00005, popmax ≤ 0.0005; ptvraredmg). For binary phenotypes, effect sizes were expressed as log(OR), with standard errors derived as (log(UCI) − log(LCI)) / 3.92; for quantitative phenotypes, beta coefficients and confidence intervals were used directly. Where multiple collapsing models yielded results for the same gene–phenotype pair, only the result with the lowest p-value was retained. Burden test statistics for all gene–phenotype pairs used as intercept priors in VIDRA are provided in the data repository, including Ensembl gene identifier, phenotype EFO code, effect size, standard error, p-value, and collapsing model category.

### Approximate rare variant effect on protein activity and disease

To quantify the effect of variants where QTL information was unavailable, we adopted an *in silico* approximation, collating the variant effect estimates of several tools. The *in silico* annotation of the variant effects on protein function was obtained via ENSEMBL Variant Effect Predictor v.111.0 using the following plugins: AlphaMissense, CADD, REVEL.^84,86–94^ These scores were independently scaled and normalised to have values ranging from 0 to 1, where values close to 0 indicate variants that almost completely inhibit protein activity (e.g., premature stop codon), and values close to 1 indicate variants that do not alter protein activity. VIDRA models coding variants along a loss-of-function (LoF) spectrum because current in silico predictors, including CADD, REVEL and AlphaMissense, are designed to identify deleterious variants but do not reliably distinguish between LoF and gain-of-function (GoF) effects. Directionality is instead inferred through integration with gene-level burden tests, which provide an anchor for LoF effects, and eQTL/pQTL data, which capture bidirectional changes in gene expression and protein abundance. Consequently, although coding variants are represented on a LoF spectrum, the inferred VIDRA slope reflects the overall relationship between gene activity and disease risk, including scenarios consistent with GoF mechanisms. Incorporating explicit GoF annotations as large-scale functional datasets become available represents an important direction for future development. The scaled scores are then used as relative protein function priors in the VIDRA Bayesian framework. When the *in silico* resources report large deviations in the reported scores, VIDRA generates a unique score weighted by the standard deviation of each score. Similarly, VIDRA estimates the pathogenicity probability distribution for a given variant from ClinVar pathogenicity reports, ordered from benign to pathogenic. ClinVar reports are an aggregation of the variant involvement in the phenotypes, how well their association is described and how penetrant this variant is. We order the reports, from the least damaging to the most damaging, as follows: not provided, association not found, other, benign, likely benign, low penetrance, confers sensitivity, uncertain risk allele, drug response, uncertain significance, association, affects, likely risk allele, risk factor, established risk allele, likely pathogenic, pathogenic. This derives a distribution of disease probabilities across a spectrum of variation akin to estimating an effect size of a variant on the risk of developing a disease with common variants. Additionally, ClinVar variants were filtered to exclude entries with conflicting interpretations or without assertion criteria.

#### Harmonisation of genetic evidence onto a common protein function axis

Variant classes are placed on a common dose axis through a variant source-specific calibration. For non-coding variants with eQTL or pQTL colocalisation, variant effect and its uncertainty correspond to the QTL effect size and standard error from summary statistics. For rare coding variants, and common coding GWAS variants lacking QTL colocalisation, the variant effect is approximated by an ensemble of in silico pathogenicity scores (AlphaMissense, CADD, REVEL, SIFT, PolyPhen, Blosum62, PrimateAI, GERP conservation, FoldX ΔΔG), each independently scaled to [0, 1] where 0 denotes near-complete loss of function and 1 denotes a benign variant; the standard error is set to a fixed weakly-informative prior of 0.1, reflecting the absence of a formal sampling distribution for *in-silico* predictions. Missense-specific scores (AlphaMissense, REVEL, PolyPhen, SIFT, PrimateAI, Blosum62) are available only for missense variants. When these scores are absent, variant effects are filled with the per-gene mean of the available missense values observed at other positions in the same gene, providing a gene-level background estimate of missense constraint. Loss-of-function consequences are hardcoded to 0 (maximally damaging). Where available, gene burden test statistics enter the model as intercept priors anchoring the baseline phenotypic state when protein function approaches zero; burden test standard error is derived from the confidence interval of the burden test odds ratio or beta. Heterogeneous uncertainty across these classes is propagated through five variant source-specific random effect slopes within the hierarchical model and a Student’s t-distribution likelihood (degrees of freedom ν = number of unique variants − 1), which robustly down-weights high-leverage observations regardless of evidence class.

### VIDRA Bayesian model for dose-response estimation

VIDRA follows the instrumental variable logic of Mendelian randomization, in which genetic variants are used to link variation in a molecular exposure to variation in a disease outcome. In standard two-sample MR, the causal effect is typically estimated from the ratio of the variant-to-outcome effect to the variant-to-exposure effect, or by inverse-variance weighting across multiple instruments. This framework is well-suited to settings in which instruments belong to a single evidence class and the exposure effects are measured on a comparable scale.

VIDRA extends this framework to accommodate the structure of the available genetic data, where instruments differ in both scale and uncertainty. eQTL and pQTL effects are measured on standardised continuous scales, whereas in silico pathogenicity scores are bounded between 0 and 1, and ClinVar annotations encode discrete severity categories. Applying a direct ratio-based MR estimator across these data types would therefore combine exposure effects that are not naturally commensurate. In addition, the number of instruments available for each gene–phenotype pair is often small, with a median of two variants in the real data (Fig. 2e), making standard inverse-variance weighted estimates unstable. Lastly, each evidence class carries systematically different measurement uncertainty, which a single-variance MR model cannot accommodate.

The VIDRA Bayesian model addresses these limitations within a hierarchical regression framework. For each variant, the x-axis represents a source-specific proxy for relative protein function, and the y-axis represents the corresponding disease effect size. Source-specific random-effect slopes allow each evidence class to contribute a class-specific estimate, which are then aggregated into a single hierarchical slope through shared priors. Gene burden statistics are incorporated as informative intercept priors, providing information on the phenotypic effect when inferred protein function approaches zero and helping to stabilise inference when variant-level instruments are sparse.

The likelihood is specified using a Student’s *t*-distribution, which reduces sensitivity to individual outlying instruments. This plays a role analogous to robust MR estimators in conventional analyses, such as weighted median or MR-Egger approaches, while remaining within the hierarchical Bayesian framework.

The resulting VIDRA slope is interpreted as a hierarchical MR-like estimate of the relationship between relative protein function and disease risk. It is expressed as the change in standardised disease effect size per unit change in inferred relative protein function on the synthetic [0,1] scale. Because this exposure axis is estimated by harmonising heterogeneous sources, including QTL effect sizes, in silico pathogenicity scores, and ClinVar ordinal encodings, VIDRA slopes should not be directly compared numerically with conventional MR coefficients. Such comparisons would conflate differences in exposure scaling with differences in biological effect size. Instead, calibration and interpretation of VIDRA slopes are assessed through rank-order correlation and directional concordance with orthogonal genetic estimates, as shown in Fig. 4a and Fig. 5f.

The data collected from Open Targets is aggregated, and all the variant information is harmonised in a single table, and then split into per-gene files and analysed per disease. These are used as input for the VIDRA Bayesian model. The VIDRA Bayesian model estimates the dependency of phenotypic risk on protein activity by regressing the effect of genetic variants on disease outcomes as a function of genetically inferred protein activity, explicitly accounting for the inherent heterogeneous uncertainty of the different data types. VIDRA models the relationship between variant effect and disease outcome using linear regression:

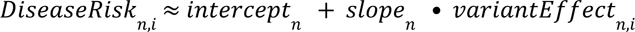

Where the *n* is the source of genetic information (e.g. ClinVar, gene burden or GWAS) and the *i* is the variant.

VIDRA models dose-response relationships between gene function and disease risk by estimating slope distributions for different sources of genetic evidence, common variants (e.g., eQTLs, pQTLs), and rare variants (e.g., ClinVar, gene burden results). Here, *variantEffect* serves as a proxy for the genetically inferred perturbation of protein activity — the genetic ‘dose’ — while *DiseaseRis*k represents the phenotypic ‘response’, directly paralleling the dose-response principle used in pharmacology. VIDRA fits a regression between relative protein function and disease outcome for each evidence source. The resulting VIDRA slope quantifies the direction and strength of the gene-phenotype relationship. To reflect biological priors, intercepts are handled differently: for eQTLs and pQTLs, we apply a weakly informative prior centred at 0 (i.e., no effect) with a standard deviation of 5; for rare variant data (e.g., ClinVar), the intercept is either inferred from gene-level burden test statistics (when available) or randomly initialized to 0, to reflect limited prior knowledge. Crucially, VIDRA’s hierarchical structure allows for modeling random effect slopes depending on the input source, aiming to improve prediction accuracy while collapsing information across all the available groups. It uses a Student’s t-distribution to model the slope estimates. This distribution is preferred over normal distribution because it is robust to outliers, capturing long-tailed uncertainty while focusing the fit on the central, high-confidence region of the slope distribution.

Posterior distributions for VIDRA slope parameters were obtained with full-rank Automatic Differentiation Variational Inference (ADVI) sampling. The variational algorithm used 20 gradient samples during optimization, and 1,000 draws were subsequently taken from the fitted variational posterior approximation. The prior distribution were weakly informative: slope assumed a null effect N(0,50) for each of the sources considered in the hierarchical model. Similarly, the intercept had a diffuse prior N(0,10). VIDRA uses Stan v2.34.1 as statistical modeling software. VIDRA Bayesian model, despite differences in the experimental designs, aims to estimate the same underlying latent variable, i.e. the dependency of phenotype risk on protein activity, which can then be used to inform drug development. While assuming linearity imposes biological limitations, since some cases may exhibit non-additive relationships between protein activity and disease, a linear model offers a straightforward interpretation of the slope. Given the limited number of variants supporting dose-response estimates, this simpler model helps prevent overfitting, reduces sensitivity to outliers, and remains more interpretable. For cases where a single variant links a gene to a phenotype, the slope estimate was calculated as previously described.^95^

In the subsequent analysis, we retained only gene-phenotype pairs with strong statistical support: a posterior probability greater than 0.7 for pairs supported solely by ClinVar and at least 0.55 for those with multiple sources of evidence. We classified the genetically derived therapeutic direction of modulation as *activators* if the dose-response relationship had a negative slope and *inhibitors* if the slope was positive. Additionally, we identified genetically predicted adverse events as phenotypes exhibiting directionality opposite to the primary disease association.

### Simulation to benchmark the VIDRA Bayesian model

We compared VIDRA to linear regression and assessed the effect of one other Bayesian model: one non-stratified regression (i.e. all variables are entered simultaneously despite their origin). We evaluated the model simulating the limited number of instrumental variables, as observed in Fig. 2e. We generated a set of defined beta slope estimates and registered how the different models could re-estimate the defined slope in various conditions. We quantified the effect of the number of variants, going from 0 (i.e. data not present in the simulated source of condition) to a maximum of 5. During the simulations, to mimic the noise and technical variability from different data sources, such as GWAS, rare coding, and ClinVar, we also introduced noise via increasing standard deviations in the *relative protein function* estimates (ranging from 0.1 to 0.5) and the regression error terms (ranging from 0.1 to 0.5).

### VIDRA Therapeutic Potential Score calculation

We implemented a stacked classifier to predict drug target approval (VIDRA Therapeutic Potential Score) by combining multiple machine learning algorithms (i.e. gradient boosting, Gaussian support vector machine (SVM), and linear SVM). This ensemble approach improves predictive accuracy by leveraging the strengths of different algorithms while reducing biases and errors that might affect individual models. The stacked classifier consists of two layers: base models and a meta-model. The base models (gradient boosting, Gaussian SVM, and linear SVM) are trained independently to predict whether a drug target will be approved, each capturing different patterns in the data to enhance robustness. The meta-model then aggregates these predictions using logistic regression. This improves generalization, preventing overfitting by combining different perspectives and ensuring a more balanced decision-making process. The ensemble model is trained on VIDRA-derived features, including the median VIDRA slope, which captures the strength of the dose-response relationship between gene function and phenotype; the number of variants, which provides a measure of data support for the gene-phenotype association; the source of variants, incorporating different genetic evidence layers (e.g., GWAS, QTLs, rare variants) to handle diverse data structures; and slope confidence, reflecting the statistical reliability of the dose-response estimate. By integrating multiple models and leveraging diverse genetic features, our stacked classifier enhances predictive performance, providing a more reliable and interpretable approach to prioritizing drug targets. Score thresholds were determined empirically by calibrating against the approved drug training set. We evaluated the enrichment of clinically approved drug targets (phase 4 drugs) relative to targets without genetic evidence as a continuous function of score threshold (Fig. 3b). A threshold of 0.7 was selected as the primary prioritisation cutoff, corresponding to a greater than 20-fold enrichment of approved drug targets. A secondary threshold of 0.6 was defined to demarcate a zone of moderate evidence, where enrichment for approved targets is statistically supported but below the stringency required for high-confidence prioritisation. Therefore, these thresholds reflect empirically defined boundaries in the score distribution where the density of clinically validated targets changes substantially (Fig. 3a, 3b). The VIDRA Therapeutic Potential Score was trained on 80% of the drug-gene-phenotype information and tested on the remaining 20%. We compared VIDRA prioritisation score and classification of the therapeutic direction of modulation to the genetic priority score (GPS).^2^ Because of the differences in the score definition and its confidence, we compared our odds ratio in the highest scoring bin (i.e. best performing) to the odds ratio reported for their best performing GPS score bin.

### Statistical analysis

To assess the statistical significance of the area under the ROC curves (AUC) for activators and inhibitors, as well as the intermediate phenotype-disease associations, we performed permutation tests with 10,000 iterations in each case. For the activator and inhibitor AUC analysis, we randomized the therapeutic direction of modulation labels and recalculated the AUC in each permutation using the scrambled dataset. Statistical significance was then determined by calculating p-values using Laplace smoothing.

Similarly, for the intermediate phenotype-disease regression, we randomized the intermediate phenotype-disease associations to estimate the level of random noise in the correlation. In each permutation, we recalculated the regression betas between intermediate phenotypes and disease, followed by p-value computation using the same approach as before. The curated list of intermediate phenotype-disease association pairs used in this analysis is provided in **Supplementary Table 3**.

The regressions for intermediate phenotype-diseases, chemical-to-genetic and adverse events were performed using robust regression with Hubers M-estimator to account for the small number of samples and compensate for the possible outlier’s effect. The algorithm used was from statsmodel v0.14.1 python package.

## Acknowledgement

We thank Lewis Ewans for his feedback interpreting the results related to the *APOE* discussion. This work was funded by Open Targets (OTAR2069). This research was funded in part, by the Wellcome Trust 220540/Z/20/A. For the purpose of Open Access, the author has applied a CC BY public copyright licence to any Author Accepted Manuscript version arising from this submission. MPA was supported by Open Targets (OTAR2065).

## Contributions

L.S. and D.C. conducted statistical analyses. L.S., and G.T. conceived the study design. L.S., D.C., Y.T. and G.T. wrote the manuscript. L.S., J.H. and J.A.L. conceived the Stan statistical model. Z.K., O.B., M.P.A., B.S., K.F., H.O., A.B., M.C.T., E.R.H., E.M.M., A.L., I.D., K.A., Y.T., D.O., J.C., N.N., and M.G., contributed to the study design and result interpretation. O.B., M.P.A., B.S., A.B., E.M.M., K.A., Y.T., and D.O. provided critical comments on the manuscript. K.A., J.C.M., N.N., and G.T. acquired funds for the research and supervised the project. All authors reviewed and approved the final version of the manuscript.

