## Supplementary figures and table legends for "Genetic dose-response modelling predicts drug mechanisms, dosing, and adverse events"

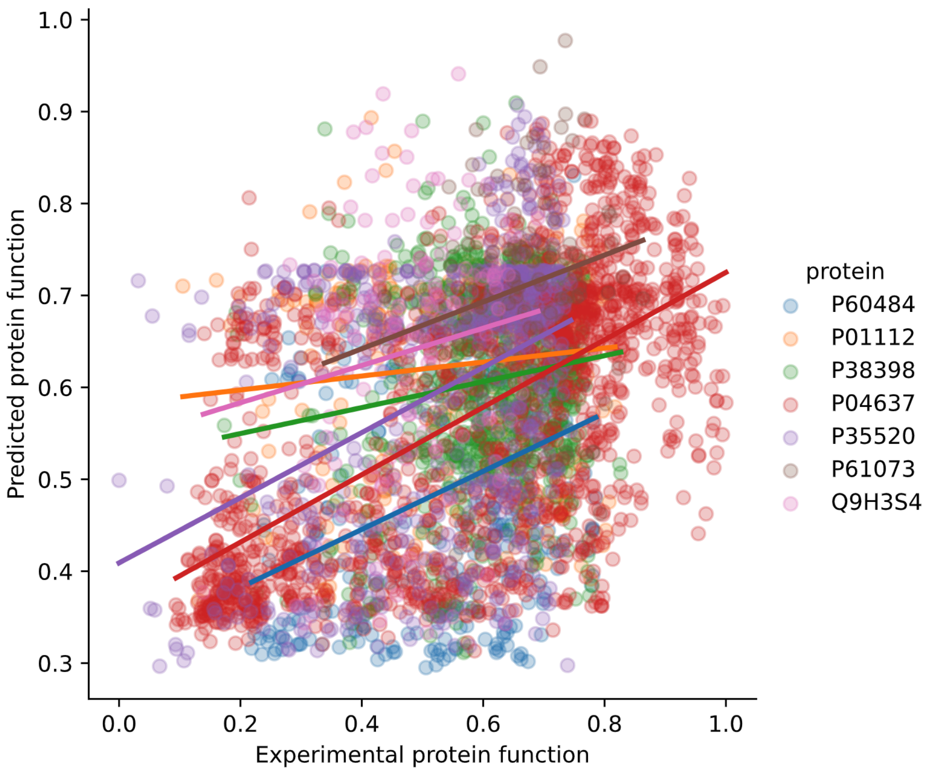


**Supplementary Figure 1 | Comparison of the experimental estimation of the variant effect on protein function to the *in-silico* predicted effect.**

Experimental predictions (x-axis) are derived from deep mutational scanning experiments and have been published in PMID 34292650. Variant estimates are scaled between 0 and 1 and transformed to a normal distribution using quantile transformation. The y-axis is an aggregated score used in the VIDRA workflow to predict the variant effect on relative protein function when QTL information is unavailable.

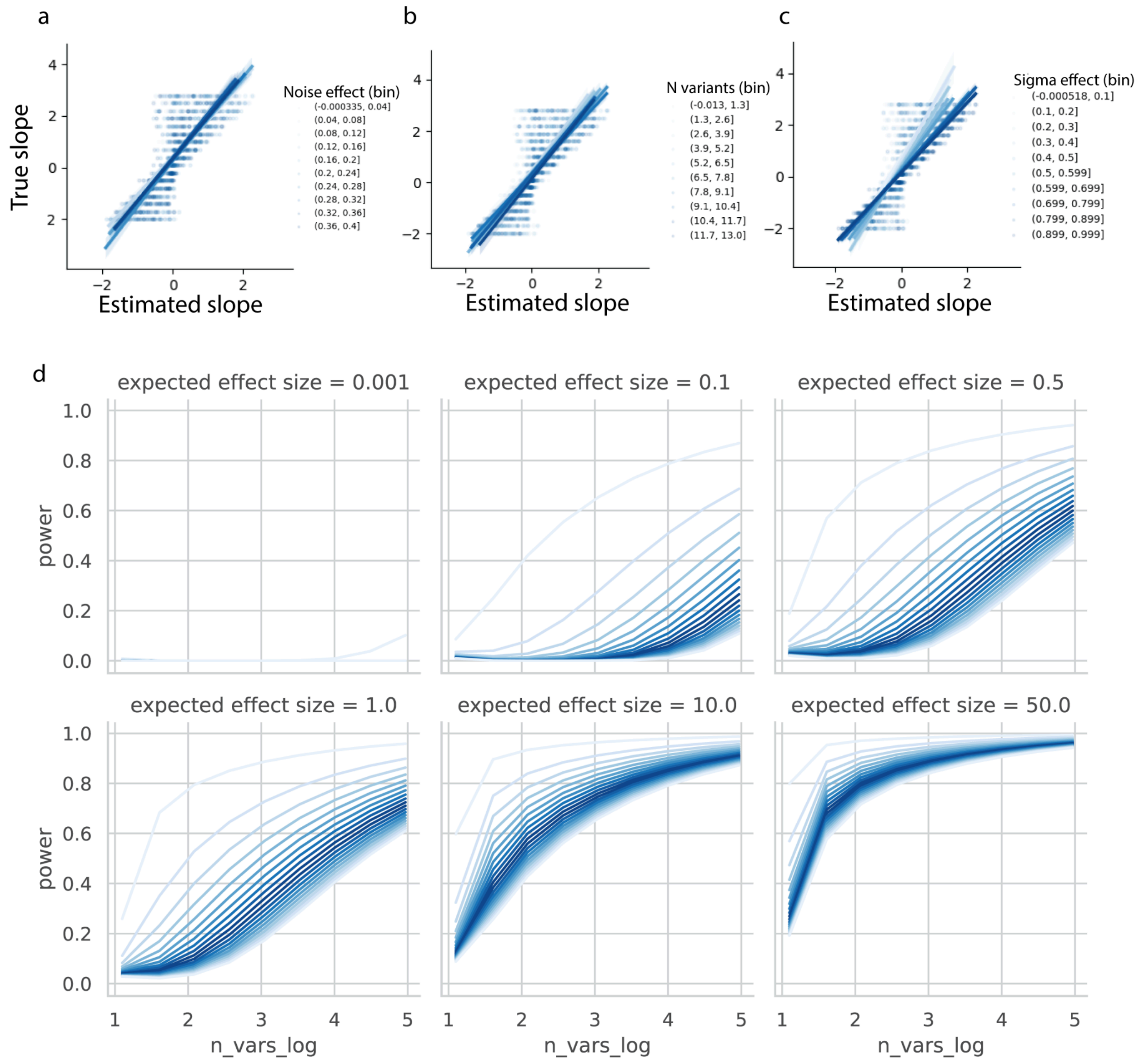


**Supplementary Figure 2 | VIDRA performance benchmark using simulated data and power calculation.**

**a–c)** VIDRA performance under varying simulation conditions. Each panel shows the relationship between the true slope values (y-axis) and the slope estimates produced by VIDRA (x-axis), across simulation iterations. Individual points represent simulation runs and are colour-coded by binned values of the simulation condition: **a)** simulation noise, **b)** number of variants (sample size), **c)** observation variance. Regression lines per bin illustrate overall trends under each condition. The plots demonstrate that slope estimation accuracy decreases with higher noise and observation variance, while larger sample sizes (more variants per gene–phenotype pair) improve the precision of the estimates. **d)** Power calculation. The x-axis (log scale) represents the number of variants used in the model, and the y-axis shows the statistical power to detect an effect. Different shades of blue correspond to different levels of residual variance (σ) used in the simulations, with lighter shades having less variance. These plots highlight how power increases with more variant observations and lower noise, guiding variant count thresholds in downstream analyses.

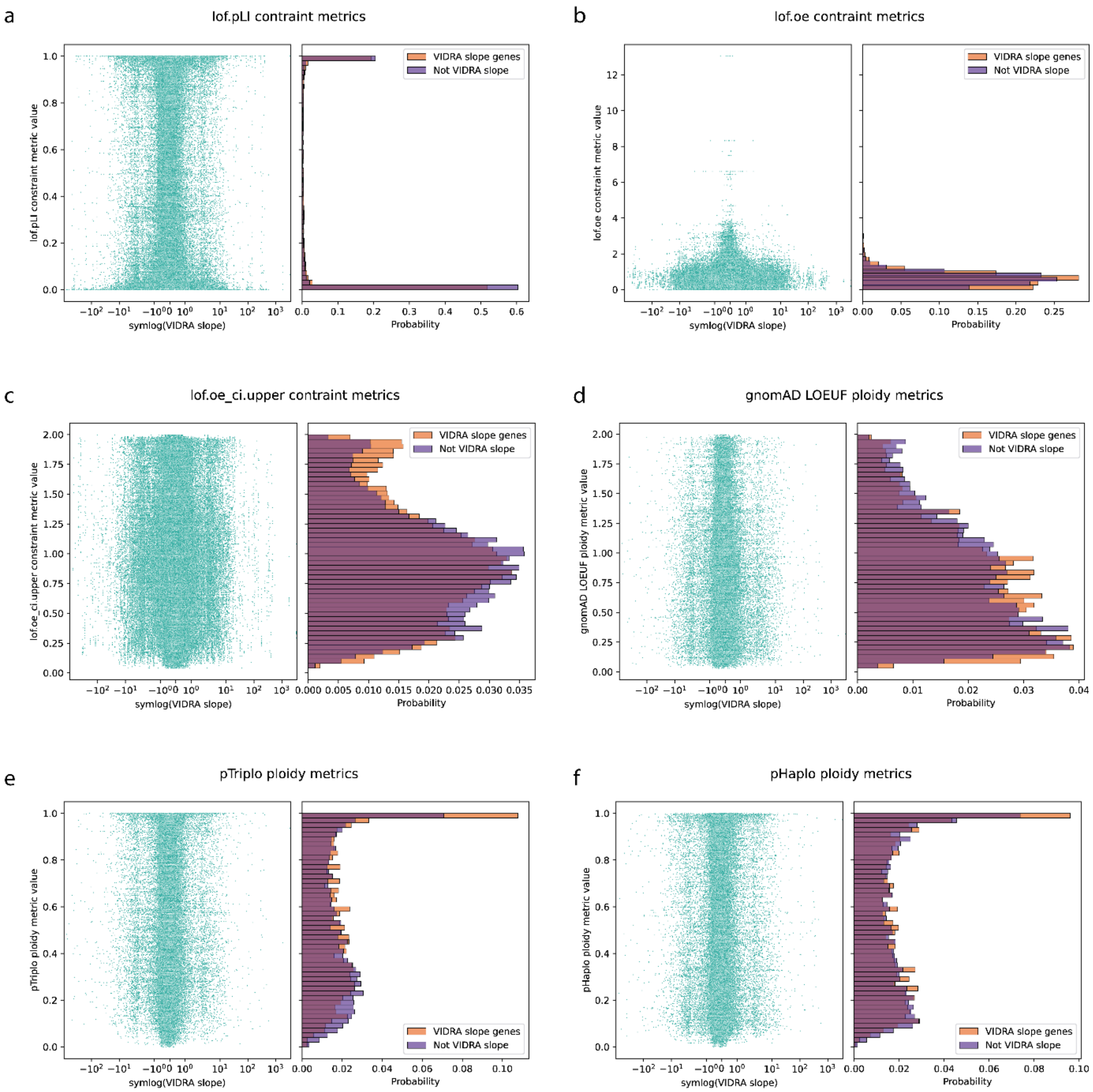


**Supplementary Figure 3 | Relationship between VIDRA slope estimates and constraint matrices.**

The left side of each panel shows the correlation between VIDRA slopes (x-axis) and pLI **(a)**, oe **(b)**, ou upper confidence interval **(c),** LOEUF **(d)**, pHaplo **(e)** and pTriplo **(f)** on the y-axis. The right side of the panels shows the distribution of constraint metrics for genes with VIDRA slopes and those without them.

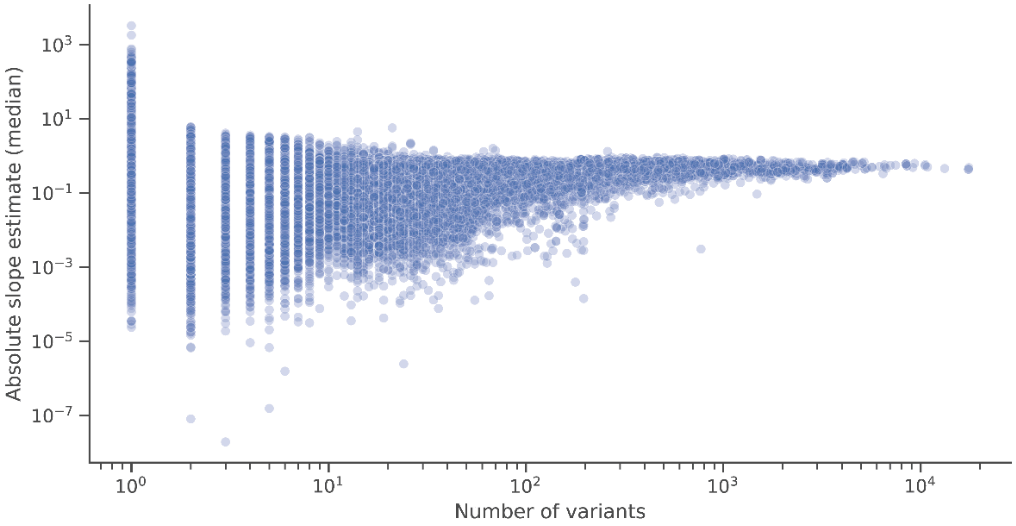


**Supplementary Figure 4 | Relationship between VIDRA slope estimates and the number of variants used in the model.**

The plots show the relationship between the number of variants included in each VIDRA model (x-axis) and the absolute value of the corresponding slope estimate (y-axis) in logarithmic scales. The analysis shows no clear positive correlation between the number of variants and the magnitude of the VIDRA slope estimate.

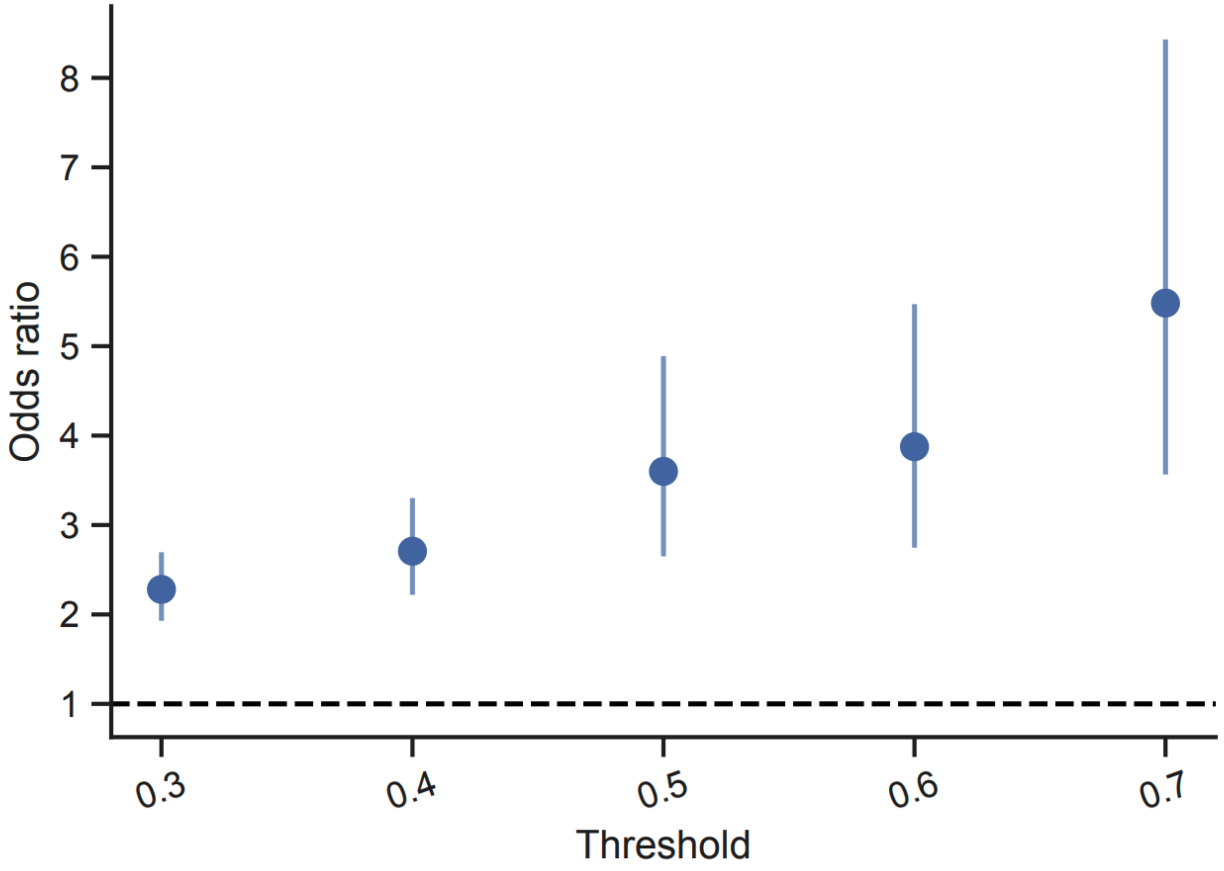


**Supplementary Figure 5 | Drug–target enrichment for VIDRA therapeutic potential scores based on gene–disease pairs.**

This figure uses a stacking classifier as described in the main Methods, but with explicit matching of gene–indication pairs to known drug target–indication pairs in the training set. Drug–indication links are propagated indirectly through the EFO/MONDO disease ontology (as in Nelson et al. 2015), and a threshold is applied to the resulting genetic-support score. The x-axis shows this therapeutic-potential score threshold; the y-axis is the odds ratio of a gene–indication pair being an approved drug target (max clinical Phase IV). Points show odds ratios with 95% confidence intervals; the dashed line (OR = 1) indicates no enrichment. Enrichment is significant at every threshold and increases with stricter cutoffs, rising from OR = 2.28 (95% CI 1.93–2.70, n = 166,204) at a threshold of 0.3 to OR = 5.48 (95% CI 3.57–8.43, n = 10,247) at 0.7.

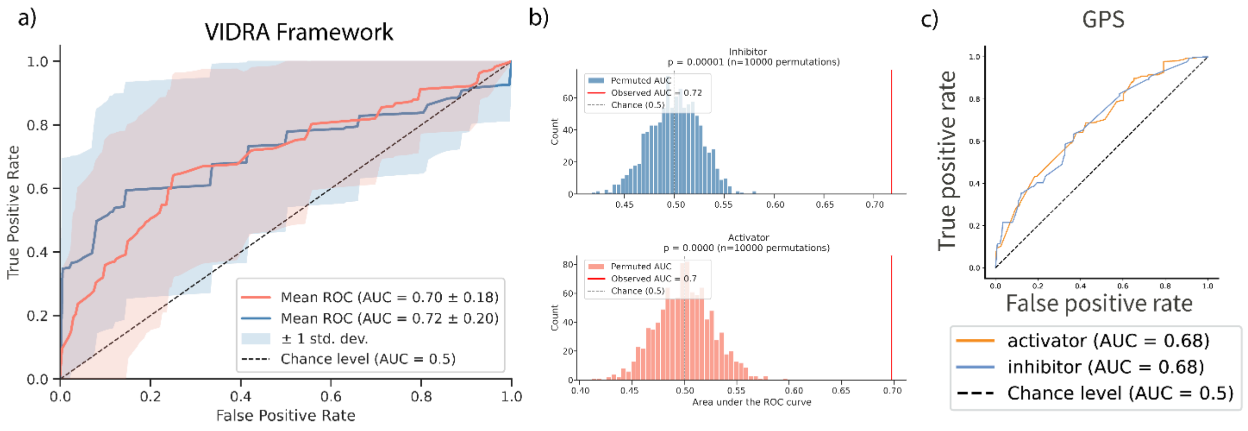


**Supplementary Figure 6 | Comparison of inferred drug directions of modulation using VIDRA framework and GPS scores.**

This figure assesses the ability of VIDRA slope directionality to infer a drug's direction of modulation, benchmarked against the GPS approach. **a)** Receiver operating characteristic (ROC) curves are used to classify directions of modulation as activators or inhibitors using the VIDRA slope direction. The y-axis shows the true positive rate (sensitivity), and the x-axis shows the false positive rate (1 − specificity). **b)** Statistical significance of the observed AUC values for activators and inhibitors, assessed by permutation testing. Direction of modulation labels were randomly shuffled across 10,000 iterations to generate a null distribution. **c)** Reference AUC values for drug directions of modulation classification based on annotations from PMID: 38172303. The diagonal dashed line in each ROC plot represents the expected performance of a random classifier. These results demonstrate that VIDRA more accurately predicts drug direction of modulation compared to GPS, particularly for distinguishing between activating and inhibitory interventions.

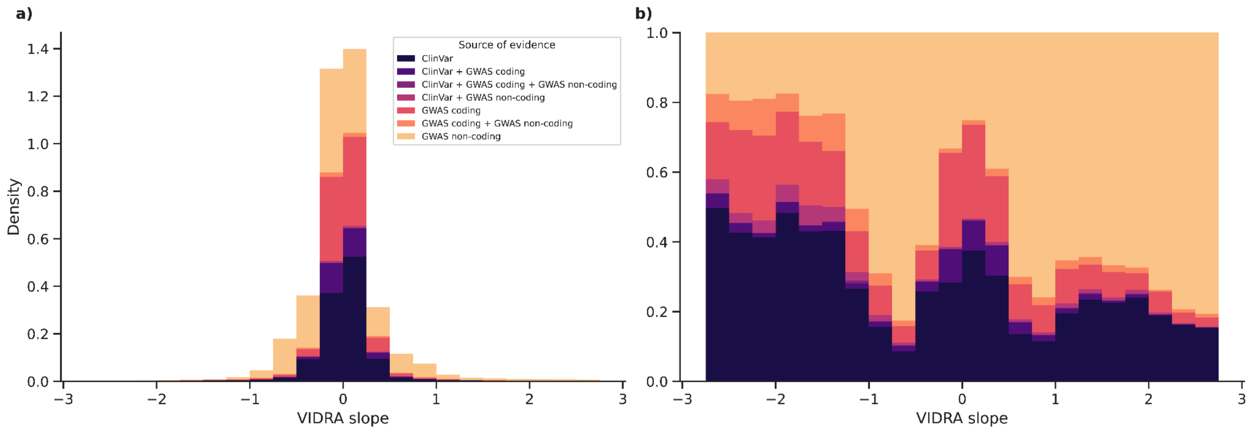


**Supplementary Figure 7 | Distribution of VIDRA slopes by variant data source.**

The plots show the distribution of VIDRA slopes, binned along the x-axis and colour-coded by the source of the variants used in each slope calculation (e.g. GWAS, ClinVar). **a)** Histogram showing the density of gene–phenotype observations across VIDRA slope bins. The height of each bar reflects the total number of observations per bin, and colours indicate the proportion contributed by each variant source. **b)** The same histogram is shown, but each bar is normalised to a total height of 1, allowing better visual comparison of variant source composition within each VIDRA slope bin. This highlights how different sources contribute across the VIDRA slope spectrum.

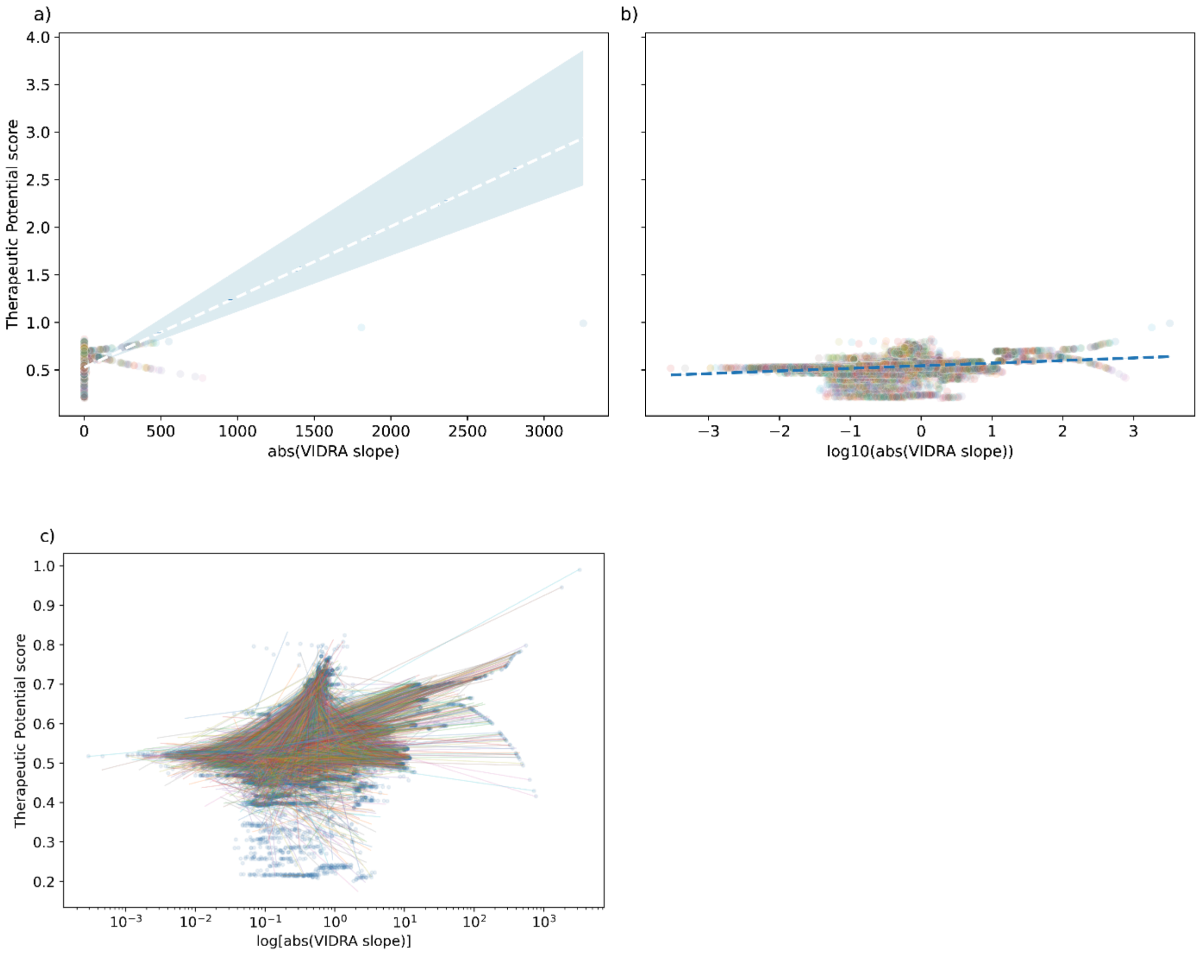


**Supplementary Figure 8 | Correlation between VIDRA slope and Therapeutic Potential score.**

**a)** The correlation between absolute VIDRA slope (x-axis) and Therapeutic Potential score (y-axis). Each dot corresponds to a gene, and they are colour-coded according to the respective associated phenotypes. The dotted blue line corresponds to the robust regression estimated coefficient representing the relationship between VIDRA slope and Therapeutic Potential score, and the blue shades are the 95% confidence interval. **b)** like a) but the x-axis has been converted to log scale to appreciate close-to-zero values better. **c)** The correlation between absolute VIDRA slope (x-axis) and Therapeutic Potential score (y-axis); each dot is a gene, and the regression estimated coefficient representing the relationship between VIDRA slope and Therapeutic Potential score is colour-coded by phenotype.

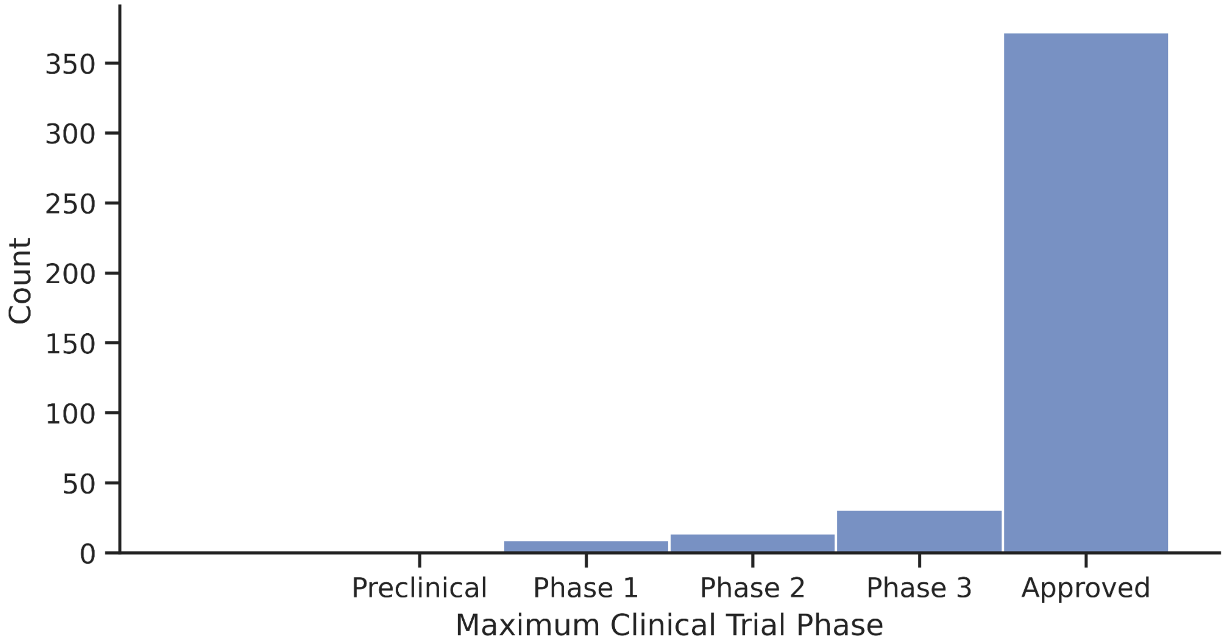


**Supplementary Figure 9 | Distribution of the maximum clinical trial phase (Open Targets v24.03) reported for genes with a Therapeutic Potential score ≥ 0.7.**

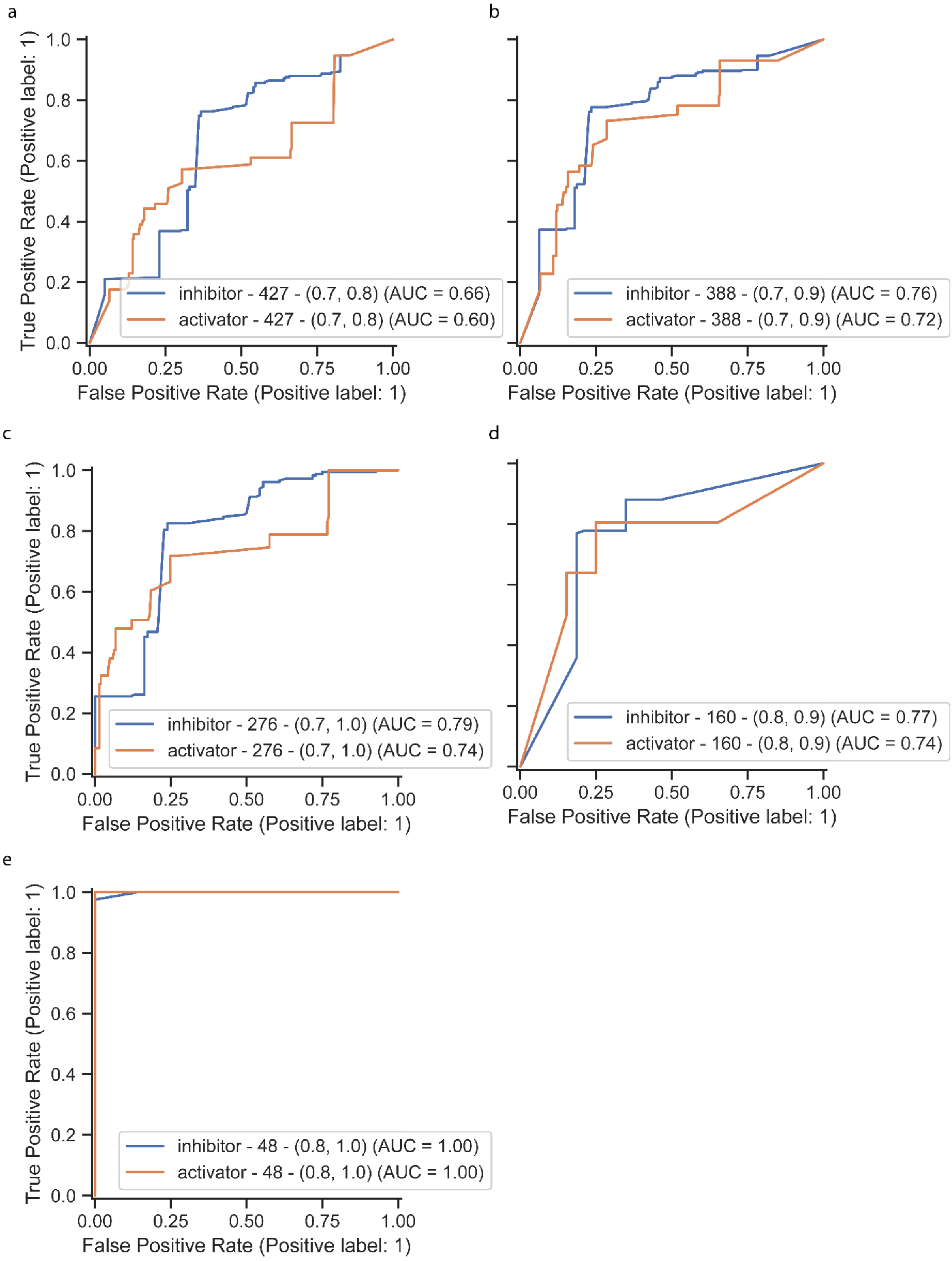

a
True Positive Rate (Positive label: 1)
False Positive Rate (Positive label: 1)
inhibitor - 427 - (0.7, 0.8) (AUC = 0.66)
activator - 427 - (0.7, 0.8) (AUC = 0.60)
b
False Positive Rate (Positive label: 1)
inhibitor - 388 - (0.7, 0.9) (AUC = 0.76)
activator - 388 - (0.7, 0.9) (AUC = 0.72)
c
True Positive Rate (Positive label: 1)
False Positive Rate (Positive label: 1)
inhibitor - 276 - (0.7, 1.0) (AUC = 0.79)
activator - 276 - (0.7, 1.0) (AUC = 0.74)
d
False Positive Rate (Positive label: 1)
inhibitor - 160 - (0.8, 0.9) (AUC = 0.77)
activator - 160 - (0.8, 0.9) (AUC = 0.74)
e
True Positive Rate (Positive label: 1)
False Positive Rate (Positive label: 1)
inhibitor - 48 - (0.8, 1.0) (AUC = 1.00)
activator - 48 - (0.8, 1.0) (AUC = 1.00)

**Supplementary Figure 10 | VIDRA slope magnitude improves classification of drug mechanism of action at more stringent thresholds.** ROC curves evaluating the ability of VIDRA slope sign to classify the direction of drug modulation (inhibitor versus activator) across increasing VIDRA slope magnitude thresholds. Each panel retains only gene–phenotype pairs whose absolute VIDRA slope exceeds the indicated threshold, with the number of pairs retained shown in each panel legend. As the slope magnitude threshold increases from 0.7 (n = 427) to 0.8 (n = 160 and n = 48 across the two highest-threshold panels), the AUC for both inhibitor and activator classification improves consistently, rising from 0.66 and 0.60 respectively at the most lenient threshold to 0.77 and 0.74 at an intermediate threshold, and reaching 1.00 for both classes at the most stringent threshold (|slope| > 0.8, n = 48).

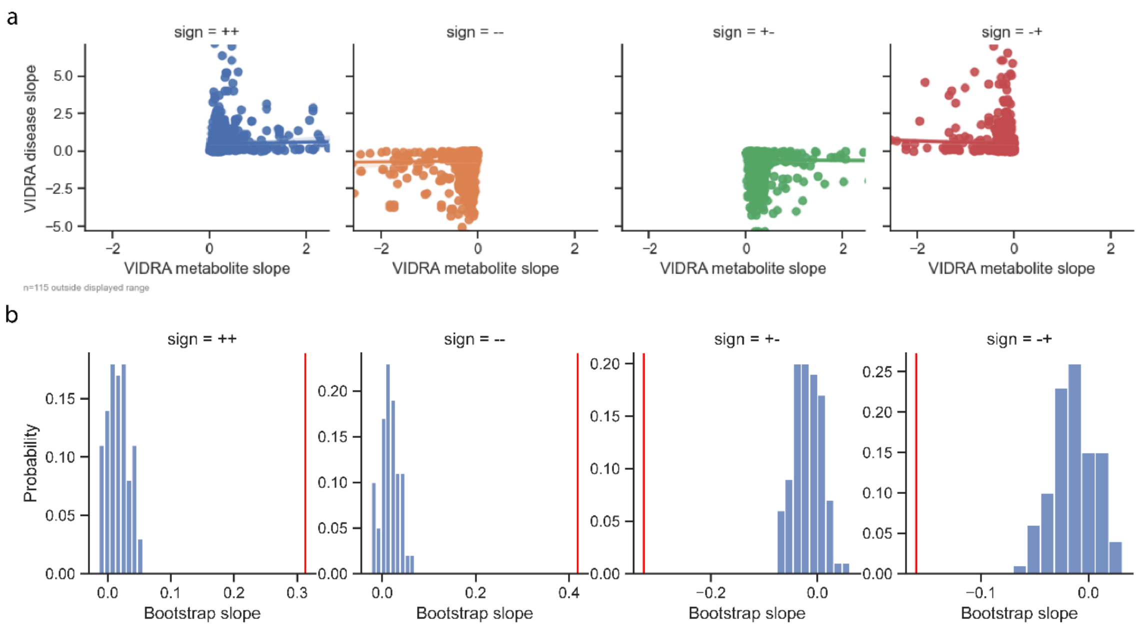


**Supplementary Figure 11 | Correlation between curated intermediate phenotypes and disease phenotypes.**

This figure builds on the analysis presented in **Fig. 4**, using the curated list of intermediate phenotype–disease pairs from **Supplementary Table 3**.

**a)** Scatterplots showing the correlation between VIDRA slope estimates for intermediate phenotypes (x-axis) and disease phenotypes (y-axis). Pairs are categorised into four quadrants based on the directionality of their slopes: both positive, both negative, intermediate positive with disease negative, and intermediate negative with disease positive. **b)** Statistical significance of the observed correlations, assessed using 1,000 permutations of the intermediate phenotype–disease pairings. These permutation tests were used to generate null distributions and calculate p-values for each directional category.

**Supplementary Table 1 | VIDRA slope estimates and posterior probabilities for gene–phenotype pairs.**
This table lists gene–phenotype pairs that passed the inclusion threshold for analysis. For each pair, the VIDRA slope estimate is provided, representing the direction and magnitude of the inferred dose-response relationship, alongside the posterior probability supporting the model fit.

**Supplementary Table 2 | Therapeutic Potential scores and corresponding clinical development stages.**
This table contains Therapeutic Potential scores for gene–disease pairs, reflecting their inferred likelihood of therapeutic success. For each pair, the highest clinical development phase achieved by a drug targeting the same gene–disease indication is also reported.

**Supplementary Table 3 | Manually curated intermediate phenotype–disease pairs.**
This table presents the curated set of intermediate phenotypes and disease outcomes used in the analysis shown in Fig. 5 and Supplementary Fig. 8. These mappings were selected based on known or hypothesised biological relevance.

**Supplementary Table 4 | Spearman correlations between VIDRA slope estimates for biomarker–disease pairs.** This table reports the pairwise Spearman correlation coefficients between VIDRA slope estimates for selected intermediate biomarker phenotypes and disease outcomes. For each biomarker–disease pair, the table provides the biomarker and disease identifiers (EFO, MONDO, Orphanet, or HP ontology terms), their respective categories, the Spearman ρ coefficient, the associated p-value, and the set of shared genes (Ensembl identifiers and gene symbols) contributing to the correlation. The table covers 863 biomarker–disease pairs across a broad range of biomarker and disease categories.

**Supplementary Table 5 | Curated clinical trial data and drug effect estimates across concentrations.**
This table summarises a manually curated set of clinical trials relevant to specific gene–disease associations. For each trial, the estimated treatment effect is shown across the reported range of drug concentrations. Clinical phenotypes have been manually mapped to corresponding genetically associated traits, enabling comparison with VIDRA slope estimates.
